# Association of Modified Cardiometabolic Index with Cardiovascular-Kidney-Metabolic Syndrome Staging, All-Cause Mortality, and Cardiovascular Mortality: A Population-Based Cohort Study

**DOI:** 10.64898/2026.03.30.26349722

**Authors:** Yuqi Qin, Yibo Yan

**Author notes:** Corresponding Author: Dr. Yibo Yan.

## Abstract

**Objective:** To investigate the association of the modified cardiometabolic index (MCMI) with cardiovascular-kidney-metabolic (CKM) syndrome staging, all-cause and cardiovascular mortality, and compare its predictive performance with traditional indices.

**Methods:** This prospective cohort study included 5,189 adults with CKM syndrome (stages 0–4) from NHANES 1999–2018 (median follow-up 10.4 years). Associations were assessed using polynomial/ordinal logistic regression, Cox models, and restricted cubic splines. Mediation analysis explored diabetes’ role. Competing risks (Fine-Gray), E-values, and sensitivity analyses ensured robustness. Predictive performance was compared using C-index and AUC.

**Results:** MCMI showed a “decelerating increase” nonlinear association with CKM staging (adjusted OR=3.90, 95%CI: 3.38–4.50). For all-cause mortality, MCMI>3.5 exhibited a threshold effect (Q4 vs Q1: HR=1.412, 1.046–1.907); RCS curves identified MCMI<3.5 as a safety interval. For cardiovascular mortality, MCMI showed a fluctuating nonlinear pattern with low-risk (3.0–3.5) and high-risk (<2.5 or >4.0) intervals. Diabetes mediated 45.5% of MCMI-cardiovascular mortality risk (total HR=1.374, indirect HR=1.141). Competing risks revealed substantial underestimation of true effects (Q4 vs Q1 sHR=3.25, trend P<0.001). MCMI remained independently associated with all-cause mortality after extensive adjustments (HR=1.22, 1.05–1.40); E-values (1.73/1.29) indicated robustness. MCMI demonstrated superior predictive performance over CMI and TyG (mean AUC difference 0.0243).

**Conclusions:** MCMI is an independent predictor of CKM progression and mortality. Its cardiovascular mortality risk is predominantly mediated by diabetes. MCMI>3.5 may serve as a clinical cut-off, outperforming traditional metabolic indices for CKM risk stratification.

## 1. Introduction

Type 2 diabetes mellitus, cardiovascular disease (CVD), and chronic kidney disease (CKD) are core disease categories currently threatening global public health. As highly prevalent non-communicable diseases, they have become key drivers of global morbidity and mortality.

Numerous studies have confirmed a high comorbidity rate among these three conditions(1). To elucidate the complex interrelationship mechanisms, the American Heart Association (AHA) first proposed the unified concept of “cardiovascular-kidney-metabolic (CKM) syndrome” in October 2023. This innovative classification emphasizes the close connections and pathophysiological interactions among metabolic abnormalities, cardiovascular lesions, and kidney injury. It is specifically defined as health impairment resulting from the interplay of obesity, diabetes mellitus, CKD, and CVD (including heart failure, atrial fibrillation, coronary heart disease, stroke, and peripheral artery disease) (2).

The pathophysiological mechanisms of cardiovascular disease, chronic kidney disease, and metabolic syndrome are all closely associated with inflammation, immune responses, and nutritional status(3). For instance, inflammation and oxidative stress act synergistically to promote atherosclerosis, exacerbate insulin resistance, and activate immune cells within the kidney to release inflammatory mediators that damage renal tubules and glomeruli(4). Overnutrition, particularly high-sugar and high-fat diets, leads to visceral fat accumulation, activating immune cells such as macrophages to infiltrate adipose tissue, forming a “chronic inflammatory microenvironment in adipose tissue.” This inflammatory state directly inhibits insulin signaling pathways (e.g., the PI3K/Akt pathway), leading to insulin resistance, which in turn further promotes lipolysis and the release of inflammatory factors, creating a vicious cycle(5).

The modified cardiometabolic index (MCMI) is a novel composite indicator developed based on the cardiometabolic index (CMI) and the triglyceride-glucose (TyG) index. It integrates triglycerides, fasting plasma glucose, high-density lipoprotein cholesterol, waist circumference, and height, providing a more comprehensive depiction of an individual’s metabolic profile and simultaneously reflecting the dyslipidemia and visceral obesity associated with insulin resistance(6). In the field of cardiovascular disease, prospective studies have shown a non-linear association between MCMI and CVD risk, demonstrating stable predictive ability for cardiovascular events(7). Furthermore, MCMI exhibits significantly superior long-term predictive value for the incidence of type 2 diabetes compared to the TyG index, particularly prominent in subgroups such as women and younger populations(8). Elevated MCMI is also significantly associated with an increased risk of sarcopenia(9). However, despite the favorable predictive value of MCMI for single metabolic or cardiovascular endpoints, the prognostic significance of this comprehensive index in the complex patient population with “cardiovascular-kidney-metabolic” multimorbidity remains a research gap. Specifically, whether MCMI can effectively predict disease progression, all-cause mortality, and cardiovascular-specific mortality in patients with CKM syndrome still needs elucidation.

Therefore, this study aims to apply the modified cardiometabolic index to the newly defined population with CKM syndrome and systematically evaluate its predictive efficacy for patient disease risk stratification, all-cause mortality, and cardiovascular mortality. We propose the core hypothesis: In patients with CKM syndrome, higher MCMI levels are independently associated with higher all-cause and cardiovascular disease mortality, and its predictive value is superior to traditional metabolic indicators. To test this hypothesis, this study will innovatively utilize the large-scale, nationally representative National Health and Nutrition Examination Survey (NHANES) database to deeply explore the association between MCMI and clinical outcomes in CKM patients, aiming to provide new evidence-based foundations for early risk identification and precision management of this high-risk population.

## 2. Methods

### 2.1 Study participants

The National Health and Nutrition Examination Survey (NHANES) is a nationally representative survey designed to collect data on the health and nutritional status of the non-institutionalized civilian population in the United States. This study utilized de-identified data from the NHANES database. Therefore, it was exempt from institutional review board approval and the requirement for informed consent. This study was conducted in accordance with the principles outlined in the Declaration of Helsinki. By employing a complex, stratified, multistage probability sampling design, NHANES ensures that its sample is nationally representative(10). Detailed information regarding the NHANES study design and data is available through its official website (www.cdc.gov/nchs/nhanes/).

This study utilized data from 10 NHANES cycles spanning 1999 to 2018. Participants who were pregnant or for whom the 10-year cardiovascular disease (CVD) risk could not be calculated using the baseline Predictable Risk of Events (PREVENT) equation were initially excluded(11).

Additionally, we excluded participants with unavailable follow-up data or missing covariate data, including statin use and poverty-to-income ratio (PIR). Finally, participants lacking data on MCMI or its components were excluded (Figure 1). The final analytical sample comprised 5,189 participants aged 30 to 79 years. This cohort of 5,189 participants was followed up, with follow-up durations ranging from 0.08 to 20.75 years and a median follow-up of 10.4 years.

**Figure 1.**
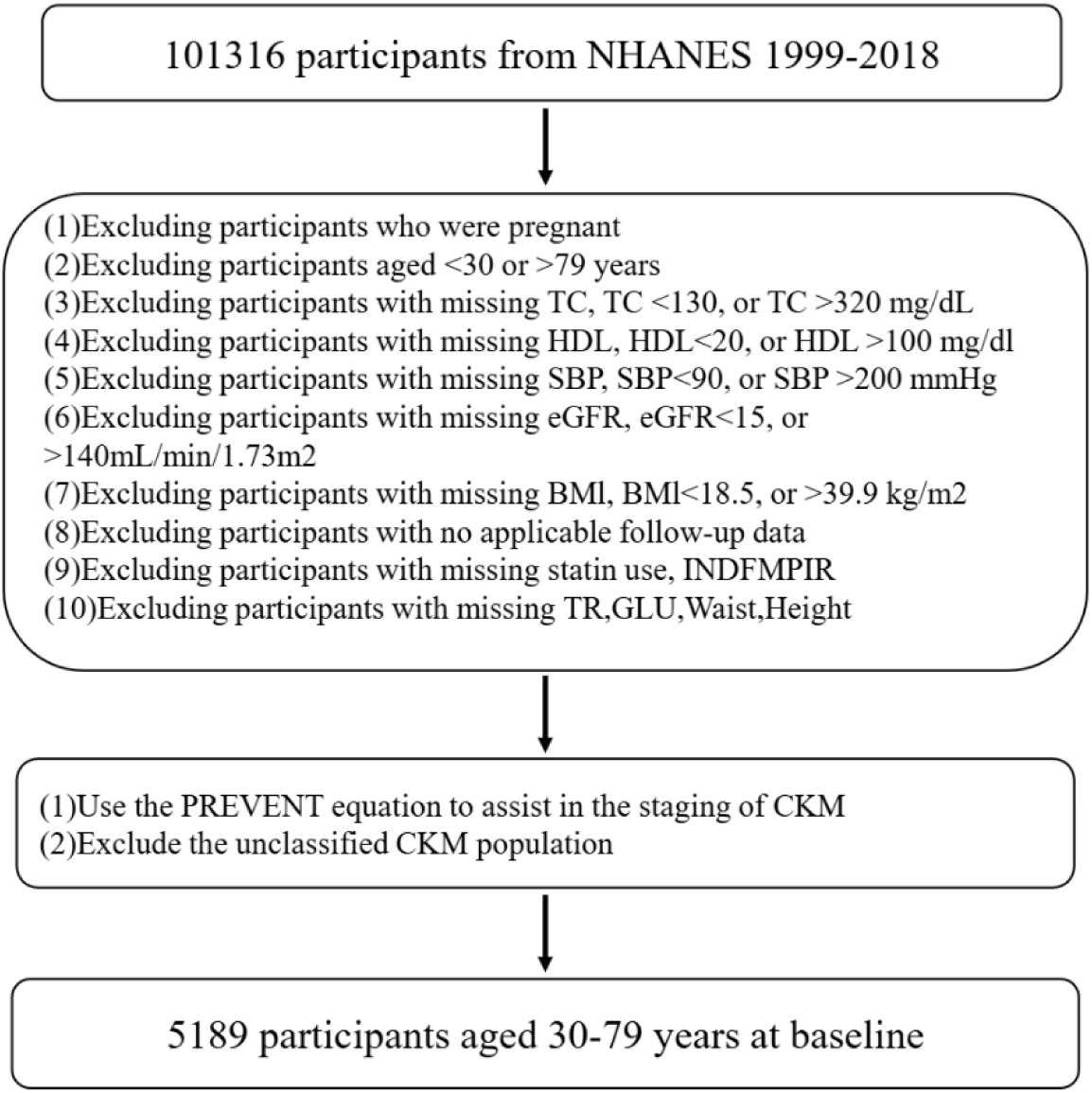
Flowchart of the study population

### 2.2 Definitions of the Modified Cardiometabolic Index, Triglyceride-Glucose Index, and Cardiometabolic Index

The TyG index was calculated as ln [TG (mg/dL) × FPG (mg/dL) / 2](12). The CMI was calculated as TG (mg/dL) / HDL-C (mg/dL) × WC (cm) / HT (cm)(13). The MCMI was calculated as ln [TG (mg/dL) × FPG (mg/dL) / HDL-C (mg/dL)] × WC (cm) / HT (cm) (6).

### 2.3 Definition of CKM stage

CKM syndrome includes individuals who are at risk of developing cardiovascular disease (CVD) as well as those with pre-existing CVD2. The stages are defined as follows:

CKM Stage 0: Individuals without overweight/obesity, metabolic risk factors (hypertriglyceridemia, hypertension, diabetes, metabolic syndrome), chronic kidney disease (CKD), or subclinical/clinical CVD.

CKM Stage 1: Individuals with overweight/obesity, abdominal obesity, or adipose tissue dysfunction, but without other metabolic risk factors, CKD, or subclinical/clinical CVD. This stage is characterized by:

(1) Body mass index (BMI) ≥25 kg/m²;
(2) Waist circumference (WC) is ≥102 cm for males or ≥88 cm for females;
(3) Fasting blood glucose (FBG) concentrations range from 100 to 124 mg/dL, or hemoglobin A1c (HbA1c) levels are in the range of 5.7% to 6.4%.

CKM Stage 2: Individuals with metabolic risk factors (including hypertriglyceridemia, hypertension, diabetes mellitus, and metabolic syndrome) or those in moderate-to-high-risk stages of chronic kidney disease (CKD) (as specified by the KDIGO criteria based on estimated glomerular filtration rate [eGFR] and urine albumin-to-creatinine ratio [UACR]). The eGFR was computed using the 2021 Chronic Kidney Disease Epidemiology Collaboration (CKD-EPI) creatinine equation, which excludes coefficients related to race and ethnicity(14).

CKM Stage 3: Individuals with subclinical CVD or its risk equivalents: high predicted 10-year CVD risk or very-high-risk KDIGO CKD stage.

(1) High 10-year CVD risk was defined as a risk ≥20%, determined using the basic Predicting Risk of CVD Events (PREVENT) equations.
(2) Very-high-risk CKD per KDIGO classification was defined as UACR ≥300 mg/g, eGFR ≤ 45–59 mL/min/1.73 m², UACR ≥30 mg/g with eGFR ≤30–44 mL/min/1.73 m², or eGFR ≤29 mL/min/1.73 m².

CKM Stage 4: Individuals with clinical CVD (self-reported diagnosis including heart failure, coronary heart disease, angina, myocardial infarction, or stroke).

### 2.4 Definition of clinical outcomes

Mortality data were retrieved from the NHANES Public-Use Linked Mortality File up to December 31, 2019. This file employed probabilistic matching methods to connect participants with the National Death Index (NDI) administered by the National Center for Health Statistics (NCHS). The causes of death were identified in accordance with the International Classification of Diseases, Tenth Revision (ICD-10), and the specified mortality outcomes underwent reclassification. All-cause mortality was defined as death resulting from any cause (ICD-10 codes: A00-Z99). Cardiovascular mortality included deaths caused by heart disease (ICD-10: I00-I09, I11, I13, I20-I51) and cerebrovascular disease (ICD-10: I60-I69).

### 2.5 Covariate assessment

We included various covariates that might influence the outcomes. Age, sex, race/ethnicity, family poverty-to-income ratio, smoking status, alcohol consumption status, medication use, and disease conditions were collected via standardized questionnaires administered during household interviews. Body mass index (BMI), waist circumference, and blood pressure were obtained through physical examinations conducted at mobile examination centers. Laboratory indicators used included C-reactive protein, neutrophil count, lymphocyte count, fasting plasma glucose, low-density lipoprotein cholesterol, high-density lipoprotein cholesterol, glycated hemoglobin (HbA1c), total cholesterol (TC), triglycerides (TG), urine albumin, creatinine (Cr), and estimated glomerular filtration rate (eGFR). Systolic and diastolic blood pressure were calculated as the mean of three measurements.

Hypertension was characterized by systolic blood pressure ≥130 mmHg, diastolic blood pressure ≥80 mmHg, a medical diagnosis of hypertension, or use of antihypertensive medication(15). Diabetes was characterized by a fasting plasma glucose level >126 mg/dL, HbA1c ≥6.5%, a medical diagnosis of diabetes, or use of insulin or glucose-lowering medications(16). Metabolic syndrome (MetS) was defined as the presence of three or more of the following: waist circumference ≥102 cm in men or ≥88 cm in women; HDL cholesterol <40 mg/dL in men or <50 mg/dL in women; triglycerides ≥150 mg/dL; elevated blood pressure (as defined above); fasting plasma glucose ≥100 mg/dL. CKD was classified as low risk, moderate-to-high risk, or very high risk according to KDIGO criteria using eGFR and UACR(17).

Alcohol consumption status was categorized as follows: heavy drinking was defined as consuming ≥3 drinks per day for women or ≥4 drinks per day for men, or binge drinking on ≥5 days per month; moderate drinking was characterized by consuming 2 drinks per day for women or 3 drinks per day for men, or binge drinking on ≥2 days per month; light drinking referred to individuals not meeting the criteria for heavy or moderate drinking; never drinking referred to individuals who consumed <12 drinks in their lifetime.

Smoking status was defined as follows: never smokers were individuals who had smoked fewer than 100 cigarettes in their lifetime; former smokers were those who had smoked at least 100 cigarettes in their lifetime but had quit smoking at the time of the survey; current smokers were those who had smoked at least 100 cigarettes in their lifetime and were still smoking at the time of the survey.

For more information on how to interpret, access, and calculate each covariate, please refer to the official NHANES analytical guidelines.

### 2.6 Statistical analysis

All statistical analyses were performed using R software, version 4.4.3. A two-sided P-value < 0.05 was considered statistically significant. The study employed weighted methods to minimize substantial fluctuations in the NHANES dataset, which utilizes a complex multistage probability sampling design to ensure accurate and representative estimates of the U.S. population. Sampling weights were adjusted according to selection probabilities and response rates; specifically, weights for the 1999–2002 cycles used wtmec 4-year * 2/10, and for the 2003–2018 cycles used wtmec 2-year * 1/10.

First, we excluded participants with missing covariate data and examined the density distribution of MCMI across different CKM stages using descriptive statistics, comparing differences between groups with analysis of variance (ANOVA). Subsequently, we performed univariable and multivariable ordinal logistic regression analyses with MCMI as a continuous variable to assess its association with CKM staging.

Next, weighted Cox regression analyses were conducted with MCMI as both a continuous variable and a categorical variable (divided into quartiles) to evaluate associations with all-cause and cardiovascular mortality in the CKM population. To address multicollinearity, we examined the variance inflation factor (VIF) of covariates before analyzing the three models and included only covariates with VIF < 5. Four regression models were developed:

Model 1: Unadjusted;

Model 2: Adjusted for demographic covariates (age, sex, poverty-to-income ratio);

Model 3: Adjusted for age, sex, poverty-to-income ratio, urine albumin-to-creatinine ratio (UACR), smoking, statin use, antihypertensive medication use, hypertension, and diabetes;

Model 4: Adjusted for age, sex, poverty-to-income ratio, UACR, smoking, statin use, antihypertensive medication use, and hypertension (excluding diabetes).

Additionally, we conducted a mediation analysis to evaluate the mediating effect of diabetes status on the association between MCMI and cardiovascular mortality risk in the CKM population. Comprehensive multicollinearity diagnostics are presented in Table 1. The proportional hazards assumption was tested using Schoenfeld residuals, and model performance was compared using the Wald test. Generalized additive models and smoothed curve fitting (restricted cubic spline [RCS] regression) were employed to characterize the dose-response relationship between MCMI and mortality. The number of knots was selected based on the minimum Akaike information criterion (AIC). Baseline characteristics for continuous variables were compared using survey-weighted linear regression, and for categorical variables using survey-weighted chi-square tests.

**Table 1.** Results of Multicollinearity Test for Covariates.

| Variable | VIF |
| --- | --- |
| Age | 1.44 |
| Sex | 1.05 |
| INDFMPIR | 1.08 |
| UACR | 1.03 |
| smoking_status | 1.17 |
| statin_use | 1.03 |
| antihypertensive_use | 1.58 |
| hypertension | 1.33 |
| diabetes | 1.17 |

\*Note: VIF: Variance Inflation Factor; VIF < 5 indicates no multicollinearity between the variable and other variables; Age: age; Sex: sex; INDFMPIR: poverty-to-income ratio; UACR: urine albumin-to-creatinine ratio; smoking\_status: smoking status; statin\_use: statin use; antihypertensive\_use: antihypertensive medication use; hypertension: hypertension; diabetes: diabetes mellitus.

To evaluate the impact of covariates on model effects and to rule out confounding, we further performed subgroup analyses and interaction tests stratified by age, sex, race/ethnicity, disease status (hypertension, diabetes, CKM syndrome), and lifestyle factors (smoking, alcohol consumption).

To ensure the robustness of our findings, a series of multidimensional sensitivity analyses were conducted for the association between MCMI and all-cause mortality risk:

1. Different grouping methods: To examine whether the categorical form of MCMI affected conclusions, we fitted models using various grouping strategies, including tertiles, quintiles, and median-based dichotomous groupings, and compared results with the quartile-based primary analysis.
2. Exclusion of specific populations: To assess potential selection bias and reverse causality, we sequentially refitted models after excluding participants with: ① follow-up duration <1 year; ② baseline cancer diagnosis; ③ baseline estimated glomerular filtration rate (eGFR) <30 mL/min/1.73 m² (severe renal insufficiency). Additionally, we separately analyzed the non-Hispanic White subgroup to preliminarily explore generalizability across different racial groups.
3. Competing risk analysis: Considering potential competing risks in mortality events (e.g., cardiovascular vs. non-cardiovascular death), we performed supplementary analyses using two approaches. First, the Fine-Gray proportional subdistribution hazards model was applied to estimate subdistribution hazard ratios (sHRs) for cardiovascular mortality across MCMI quartiles, and cumulative incidence function curves were plotted. Second, cause-specific Cox models were fitted, treating non-cardiovascular death and cardiovascular death as competing events, to assess the direct effect of MCMI on specific causes of death.
4. Model specification analysis: To evaluate the impact of model assumptions and unmeasured confounding, we conducted the following: ① Natural splines were used to model age and MCMI to examine nonlinear relationships with outcomes; ② A gradient adjustment strategy was employed, starting from a minimal set (age and sex) to a fully adjusted set, progressively incorporating socioeconomic, behavioral, and clinical covariates to observe changes in effect estimates; ③ The E-value method was applied to quantify the strength of unmeasured

confounding needed to fully explain the observed risk associations, thereby assessing robustness against potential unmeasured confounders.

Finally, we compared the predictive performance of MCMI, CMI, and TyG for mortality in the CKM population by calculating C-indices and AUC values from Cox regression models.

Continuous variables were presented as weighted means with 95% confidence intervals (CIs), and categorical variables as weighted percentages with 95% CIs. Interaction tests were used to assess statistical significance, and forest plots were generated for visualization.

## 3. Results

### 3.1 Baseline Characteristics of Participants Grouped by MCMI Quartiles

In Table 2, the study population was divided into four groups according to MCMI quartiles. Overall, as MCMI increased, participants were characterized by older age, a higher proportion of males, poorer socioeconomic status, and a greater burden of cardiometabolic risk factors. Higher MCMI groups exhibited elevated cardiometabolic disease risk, including hypertension, hyperglycemia, dyslipidemia, obesity, and a higher prevalence of chronic diseases. Of note, all-cause mortality and cardiovascular mortality among CKM patients increased with ascending MCMI.

**Table 2.** Baseline characteristics of participants grouped by MCMI quartiles.

| Variable | Q1 | Q2 | Q3 | Q4 | p |
| --- | --- | --- | --- | --- | --- |
| n | 1298 | 1297 | 1297 | 1297 |  |
| Age<br>(mean (SD)) | 50.66 (13.76) | 55.52 (13.23) | 56.32 (13.91) | 58.76 (12.76) | <0.001 |
| age_group ≥65<br>(%) | 253 (19.5) | 376 (29.0) | 443 (34.2) | 497 (38.3) | <0.001 |
| Sex = male (%) | 598 (46.1) | 742 (57.2) | 761 (58.7) | 747 (57.6) | <0.001 |
| Race (%) |  |  |  |  | <0.001 |
| 1 | 52 (4.0) | 115 (8.9) | 165 (12.7) | 196 (15.1) |  |
| 2 | 64 (4.9) | 75 (5.8) | 88 (6.8) | 118 (9.1) |  |
| 3 | 632 (48.7) | 622 (48.0) | 667 (51.4) | 701 (54.0) |  |
| 4 | 430 (33.1) | 370 (28.5) | 271 (20.9) | 209 (16.1) |  |
| 5 | 120 (9.2) | 115 (8.9) | 106 (8.2) | 73 (5.6) |  |
| INDFMPIR<br>(mean (SD)) | 3.06 (1.66) | 2.95 (1.66) | 2.80 (1.63) | 2.51 (1.60) | <0.001 |
| eGFR<br>(mean (SD)) | 116.06 (16.38) | 113.30 (18.75) | 112.85 (19.39) | 110.23 (21.04) | <0.001 |
| UACR<br>(mean (SD)) | 26.16 (200.58) | 39.10 (258.87) | 39.91 (247.28) | 83.62 (505.69) | <0.001 |
| mSBP<br>(mean (SD)) | 121.96 (18.23) | 127.13 (17.65) | 126.99 (16.77) | 129.70 (17.92) | <0.001 |
| mDBP<br>(mean (SD)) | 70.69 (11.65) | 72.54 (12.09) | 71.82 (13.11) | 72.26 (12.60) | 0.001 |
| HDL-C<br>(mean (SD)) | 66.00 (13.96) | 55.09 (12.43) | 48.47 (10.91) | 42.12 (10.27) | <0.001 |
| LDL-C<br>(mean (SD)) | 113.99 (31.22) | 123.92 (32.97) | 123.72 (34.25) | 117.23 (35.00) | <0.001 |
| TG<br>(mean (SD)) | 75.12 (29.85) | 110.75 (45.05) | 148.43 (63.83) | 228.17 (129.23) | <0.001 |
| TC<br>(mean (SD)) | 195.03 (34.48) | 201.12 (36.74) | 201.83 (38.35) | 202.34 (40.74) | <0.001 |
| HbA1c<br>(mean (SD)) | 5.42 (0.53) | 5.64 (0.69) | 5.76 (0.84) | 6.30 (1.39) | <0.001 |
| BMI<br>(mean (SD)) | 24.16 (3.26) | 27.59 (3.56) | 29.78 (3.84) | 32.61 (3.73) | <0.001 |
| WC<br>(mean (SD)) | 86.14 (8.71) | 97.15 (8.40) | 103.24 (8.94) | 111.47 (9.58) | <0.001 |
| Smoking status<br>(%) |  |  |  |  | <0.001 |
| Current smoker | 316 (24.3) | 283 (21.8) | 270 (20.8) | 288 (22.2) |  |
| Former smoker | 251 (19.3) | 318 (24.5) | 378 (29.1) | 426 (32.8) |  |
| Never smoker | 731 (56.3) | 696 (53.7) | 649 (50.0) | 583 (44.9) |  |
| Drinking status<br>(%) |  |  |  |  | 0.631 |
| Heavy drinker | 26 (2.4) | 30 (2.7) | 29 (2.6) | 27 (2.4) |  |
| Light drinker | 905 (83.0) | 883 (80.7) | 909 (81.2) | 906 (80.3) |  |
| Moderate drinker | 47 (4.3) | 43 (3.9) | 52 (4.6) | 43 (3.8) |  |
| Never drinker | 113 (10.4) | 138 (12.6) | 130 (11.6) | 152 (13.5) |  |
| Statin use (%) | 45 (3.5) | 88 (6.8) | 84 (6.5) | 108 (8.3) | <0.001 |
| Antihypertensive<br>use (%) | 309 (23.8) | 494 (38.1) | 572 (44.1) | 736 (56.7) | <0.001 |
| Hypertension (%) | 675 (52.0) | 875 (67.5) | 891 (68.7) | 991 (76.4) | <0.001 |
| Diabetes (%) | 56 (4.3) | 164 (12.6) | 253 (19.5) | 532 (41.0) | <0.001 |
| Mets (%) | 65 (5.0) | 430 (33.2) | 846 (65.2) | 1207 (93.1) | <0.001 |
| CKD risk (%) |  |  |  |  | <0.001 |
| Low risk | 1222 (94.1) | 1151 (88.7) | 1150 (88.7) | 1035 (79.8) |  |
| Moderate to<br>high risk | 63 (4.9) | 114 (8.8) | 115 (8.9) | 197 (15.2) |  |
| Very high risk | 13 (1.0) | 32 (2.5) | 32 (2.5) | 65 (5.0) |  |
| CKM stage (%) |  |  |  |  | <0.001 |
| 0 | 404 (31.1) | 15 (1.2) | 0 (0.0) | 0 (0.0) |  |
| 1 | 145 (11.2) | 200 (15.4) | 69 (5.3) | 12 (0.9) |  |
| 2 | 628 (48.4) | 859 (66.2) | 940 (72.5) | 825 (63.6) |  |
| 3 | 33 (2.5) | 76 (5.9) | 109 (8.4) | 180 (13.9) |  |
| 4 | 88 (6.8) | 147 (11.3) | 179 (13.8) | 280 (21.6) |  |
| cvd_death(%) | 26 (2.0) | 47 (3.6) | 64 (4.9) | 85 (6.6) | <0.001 |
| All-cause mortality (%) | 101 (7.8) | 150 (11.6) | 176 (13.6) | 266 (20.5) | <0.001 |

\*Note: Continuous variables are presented as mean ± standard deviation, and intergroup comparisons were performed using one-way analysis of variance (ANOVA); continuous variables not following a normal distribution are presented as median (interquartile range), and intergroup comparisons were performed using the Kruskal-Wallis H test. Categorical variables are presented as counts (percentages), and intergroup comparisons were performed using the chi-square test. Age: age; Sex: sex; Race: race/ethnicity; INDFMPIR: family poverty-to-income ratio; eGFR: estimated glomerular filtration rate; UACR: urine albumin-to-creatinine ratio; SUA: serum uric acid; mSBP: mean systolic blood pressure; mDBP: mean diastolic blood pressure; HDL-C: high-density lipoprotein cholesterol; LDL-C: low-density lipoprotein cholesterol; TG: triglycerides; TC: total cholesterol; HbA1c: glycated hemoglobin; BMI: body mass index; WC: waist circumference; Smoking status: smoking status; Drinking status: alcohol consumption status; Statin use: statin use; Antihypertensive use: antihypertensive medication use; Hypertension: hypertension; Diabetes: diabetes mellitus; Mets: metabolic syndrome; CKD: chronic kidney disease; cvd\_death: cardiovascular mortality; All-cause mortality: all-cause mortality.

### 3.2 Association Between MCMI and CKM Staging

#### 3.2.1 Distribution Characteristics of MCMI Across Different CKM Stages

The density distribution of MCMI across different CKM stages showed distinct stage-specific patterns (Figure 2). After adjustment for sampling weights, the mean MCMI exhibited a non-linear trend of initially increasing and then decreasing with advancing CKM stage: 2.19±0.28 for stage 0, increasing to 2.92±0.40 for stage 1, further increasing to 3.34±0.65 for stage 2, peaking at 3.75 ±0.76 for stage 3, and then decreasing to 3.56±0.77 for stage 4 (Table 3). The weighted sample size was largest for CKM stage 2, as this stage accounted for the highest proportion of the overall population. Furthermore, the standard deviation of MCMI increased with advancing CKM stage (from 0.28 to 0.77), suggesting greater inter-individual heterogeneity of MCMI in advanced CKM stages.

**Figure 2.**
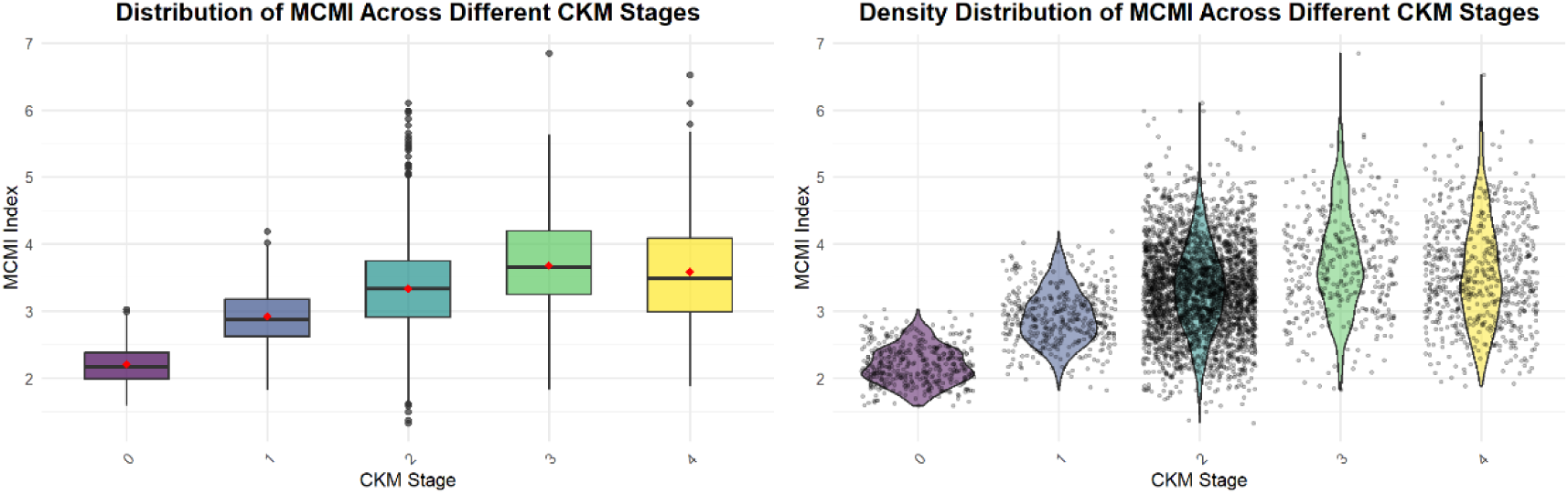
caption = “Box plots (left) display the median, quartiles, and extreme values of MCMI across different CKM stages, with red squares representing weighted mean points; violin plots (right) show the density distribution of MCMI across different CKM stages.“(The distribution range of MCMI gradually widens with advancing CKM stage, with few extreme values, and exhibits a non-linear trend of initially increasing followed by decreasing, indicating a non-linear relationship between MCMI and CKM stage.)

**Table 3.**
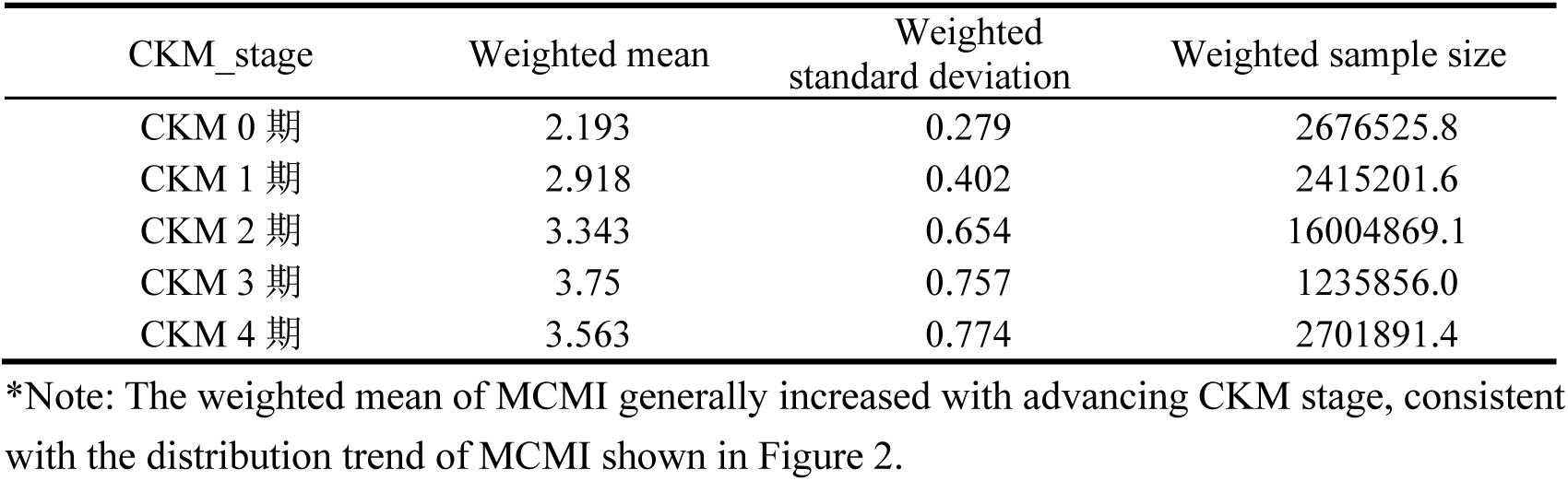
Descriptive statistics of MCMI distribution across different CKM stages.

| CKM_stage | Weighted mean | Weighted standard deviation | Weighted sample size |
| --- | --- | --- | --- |
| CKM 0 期 | 2.193 | 0.279 | 2676525.8 |
| CKM 1 期 | 2.918 | 0.402 | 2415201.6 |
| CKM 2 期 | 3.343 | 0.654 | 16004869.1 |
| CKM 3 期 | 3.75 | 0.757 | 1235856.0 |
| CKM 4 期 | 3.563 | 0.774 | 2701891.4 |

\*Note: The weighted mean of MCMI generally increased with advancing CKM stage, consistent with the distribution trend of MCMI shown in Figure 2.

#### 3.2.2 Intergroup Comparison of MCMI Across CKM Stages

After Bonferroni multiple comparison correction, except for the comparison between CKM stage 4 and CKM stage 3, which showed no statistically significant difference in MCMI (adjusted p=0.713, ns), all other pairwise comparisons between stages were highly significant (adjusted p<0.001, *) (Table 4). Specifically, the difference values for CKM stages 1-3 versus stage 0 were all negative, indicating that the adjusted mean MCMI values for these stages were lower than that for stage 0. The difference values for CKM stage 4 versus stages 1 and 2 were positive, indicating that the mean MCMI for stage 4 was higher than those for stages 1 and 2. The non-significant difference between CKM stage 4 and CKM stage 3 suggests that MCMI levels tend to stabilize in advanced CKM stages. The significant differences in MCMI across CKM stages and the observation that mean MCMI values were higher in advanced stages compared to earlier stages collectively indicate an association between CKM staging and MCMI.

**Table 4.**
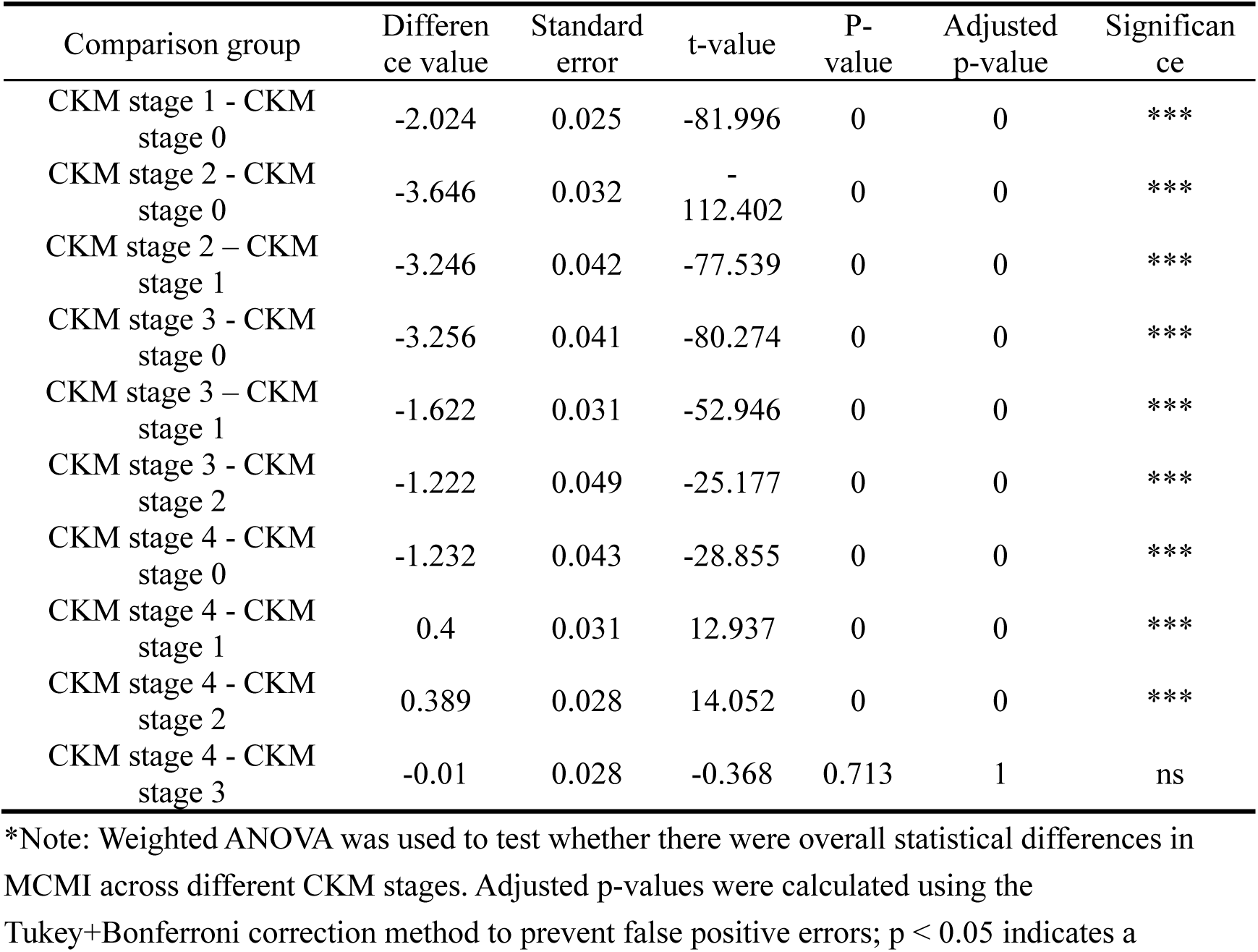
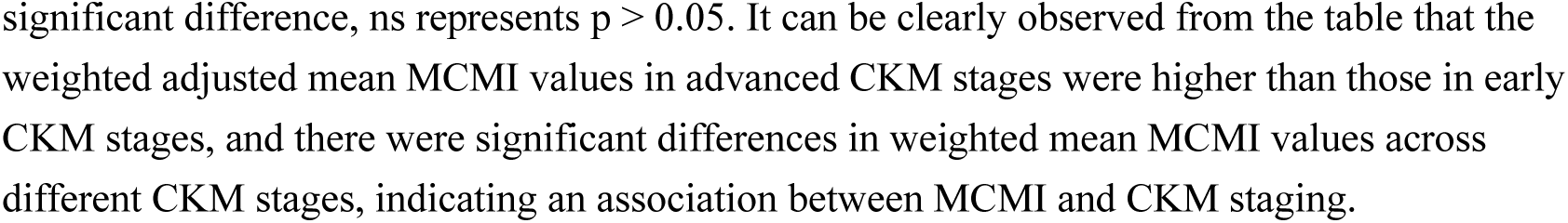
Intergroup Comparison of MCMI Distribution Across Different CKM Stages.

| Comparison group | Difference value | Standard error | t-value | P-value | Adjusted p-value | Significance |
| --- | --- | --- | --- | --- | --- | --- |
| CKM stage 1 - CKM stage 0 | -2.024 | 0.025 | -81.996 | 0 | 0 | *** |
| CKM stage 2 - CKM stage 0 | -3.646 | 0.032 | -112.402 | 0 | 0 | *** |
| CKM stage 2 - CKM stage 1 | -3.246 | 0.042 | -77.539 | 0 | 0 | *** |
| CKM stage 3 - CKM stage 0 | -3.256 | 0.041 | -80.274 | 0 | 0 | *** |
| CKM stage 3 - CKM stage 1 | -1.622 | 0.031 | -52.946 | 0 | 0 | *** |
| CKM stage 3 - CKM stage 2 | -1.222 | 0.049 | -25.177 | 0 | 0 | *** |
| CKM stage 4 - CKM stage 0 | -1.232 | 0.043 | -28.855 | 0 | 0 | *** |
| CKM stage 4 - CKM stage 1 | 0.4 | 0.031 | 12.937 | 0 | 0 | *** |
| CKM stage 4 - CKM stage 2 | 0.389 | 0.028 | 14.052 | 0 | 0 | *** |
| CKM stage 4 - CKM stage 3 | -0.01 | 0.028 | -0.368 | 0.713 | 1 | ns |

\*Note: Weighted ANOVA was used to test whether there were overall statistical differences in MCMI across different CKM stages. Adjusted p-values were calculated using the Tukey+Bonferroni correction method to prevent false positive errors; $p < 0.05$ indicates a
significant difference, ns represents $p > 0.05$ . It can be clearly observed from the table that the weighted adjusted mean MCMI values in advanced CKM stages were higher than those in early CKM stages, and there were significant differences in weighted mean MCMI values across different CKM stages, indicating an association between MCMI and CKM staging.

#### 3.2.3 Association Between MCMI and CKM Staging and the Influence of Covariates (Linear Regression)

As shown in Table 5, polynomial linear regression further revealed a non-linear positive association between MCMI and CKM staging. The model intercept was 3.3327 (p < 2e−16), indicating a significant baseline mean level of MCMI in the study population after controlling for the non-linear effects of CKM staging. The linear term (β=0.9932, p < 2e-16) indicated a fundamental increasing trend of MCMI with advancing CKM stage, while the quadratic term (β=-0.4897, p < 2e-16) suggested that this increasing trend followed a “decelerating increase” convex curve (i.e., the increase in MCMI slowed with more advanced stages). The cubic and quartic terms were not statistically significant (p > 0.05), indicating that the linear plus quadratic terms sufficiently described the association.

**Table 5.**
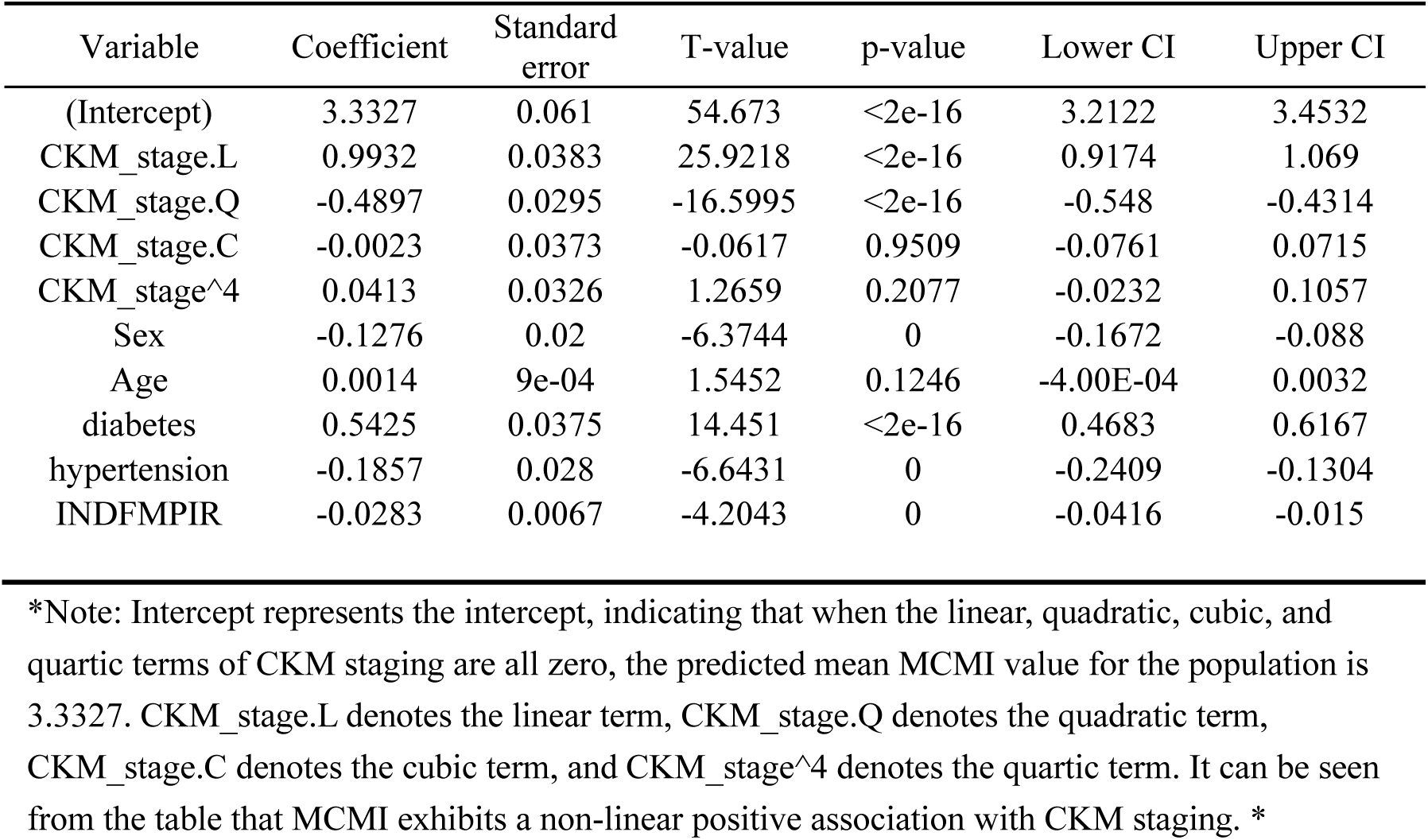
Polynomial Linear Regression Analysis of MCMI and CKM Staging.

Covariate analysis revealed that individuals with comorbid diabetes (β=0.5425, p < 2e-16) had higher MCMI values, while females (β=-0.1276, p < 0.001) had lower MCMI values. Age showed no significant effect on MCMI (p=0.1246).

#### 3.2.4 Predictive Value of MCMI for CKM Staging (Ordinal Logistic Regression)

Ordinal logistic regression analysis revealed that in the univariable model, each one-unit increase in MCMI was associated with a significant increase in the log odds of progressing to a higher CKM stage (β=1.58, p<2e-16, OR≈4.85), indicating that MCMI is a risk factor for CKM staging (Table 6).

**Table 6.**
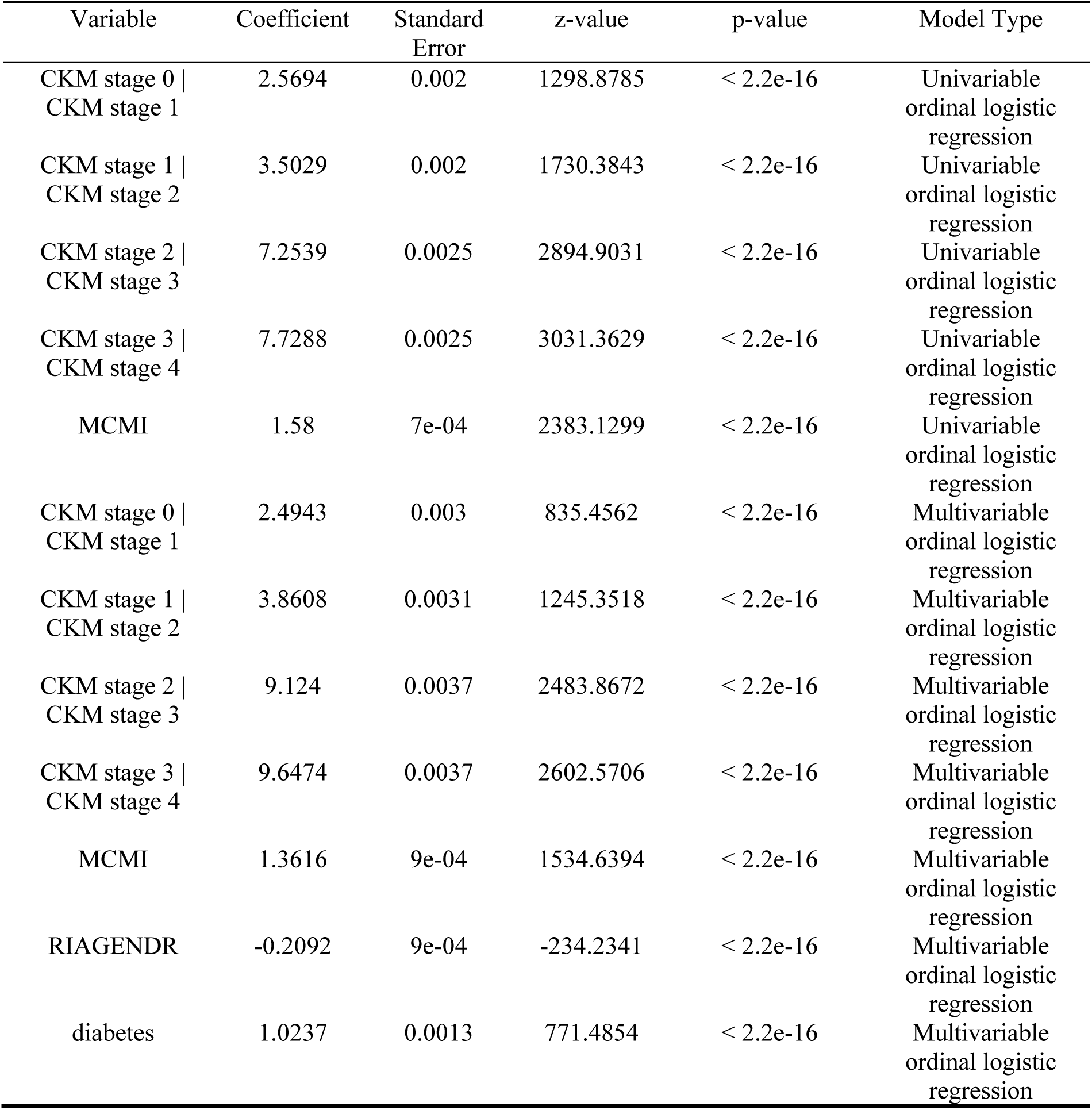
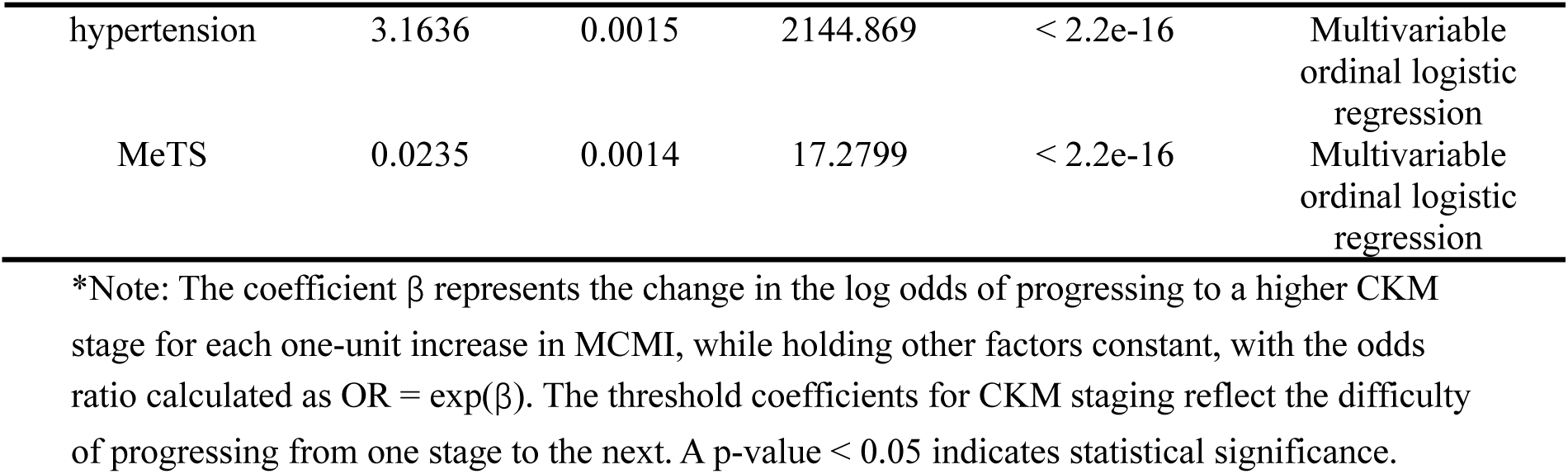
Ordinal Logistic Regression Analysis of MCMI and CKM Staging.

After adjusting for sex, diabetes, hypertension, metabolic syndrome (MeTS), and the poverty-to-income ratio (INDFMPIR), MCMI remained independently and positively associated with CKM staging (β=1.36, p<2e-16, OR≈3.90), demonstrating that MCMI is an independent risk factor for CKM stage progression. Concurrently, comorbid diabetes (OR≈2.78), hypertension (OR≈23.6), and female sex (OR≈0.81) also significantly influenced CKM staging, suggesting that these factors are independent determinants of CKM stage.

The threshold coefficients between stages (e.g., CKM stage 3 | 4 threshold = 9.6474) reflect the “difficulty” of progressing from one stage to the next. The threshold for CKM stage 3 | 4 was the highest (univariable ordinal logistic regression: 7.73; multivariable ordinal logistic regression: 9.65), indicating that progression from CKM stage 3 to stage 4 requires a higher MCMI level or exposure to more risk factors.

### 3.3 Association Between MCMI and All-Cause and Cardiovascular Mortality in the CKM Population

#### 3.3.1 Association Between MCMI as a Continuous Variable and Mortality in the CKM Population

As shown in Table 7, in the multivariable Cox proportional hazards models analyzing MCMI as a continuous variable, each one-unit increase in MCMI was associated with a 79.3% increased risk of all-cause mortality (HR=1.793, 95% CI: 1.615-1.992, p<0.001) and a significantly increased risk of cardiovascular mortality (HR=2.037, 95% CI: 1.676-2.476, p<0.001) in the unadjusted model. After adjusting for demographic variables, each one-unit increase in MCMI was associated with a 36.9% increased risk of all-cause mortality (HR=1.369, 95% CI: 1.217-1.539, p<0.001) and a 53.1% increased risk of cardiovascular mortality (HR=1.531, 95% CI: 1.228-1.907, p<0.001).

**Table 7.**
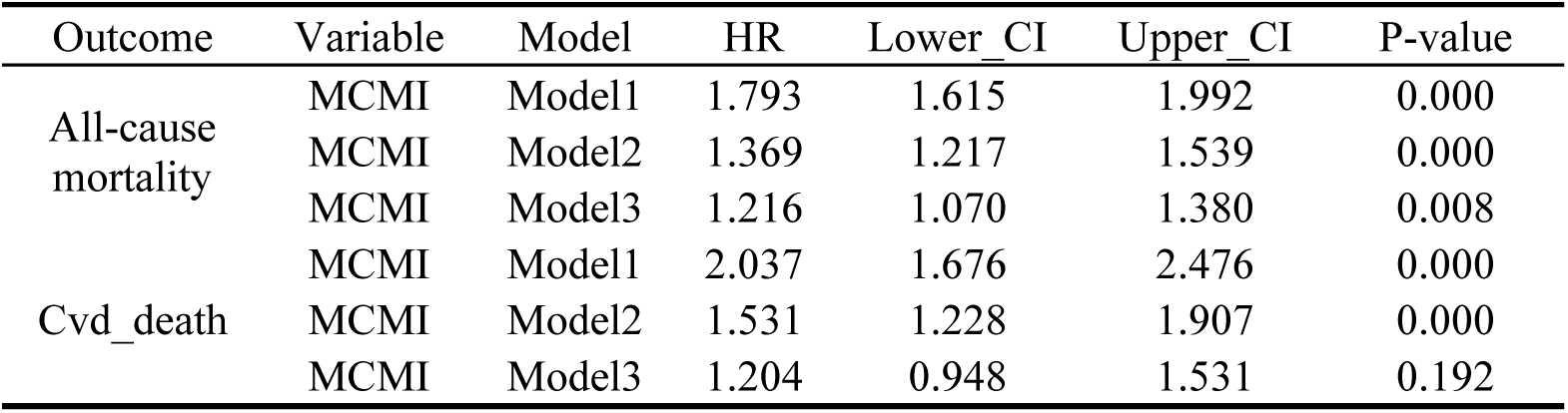
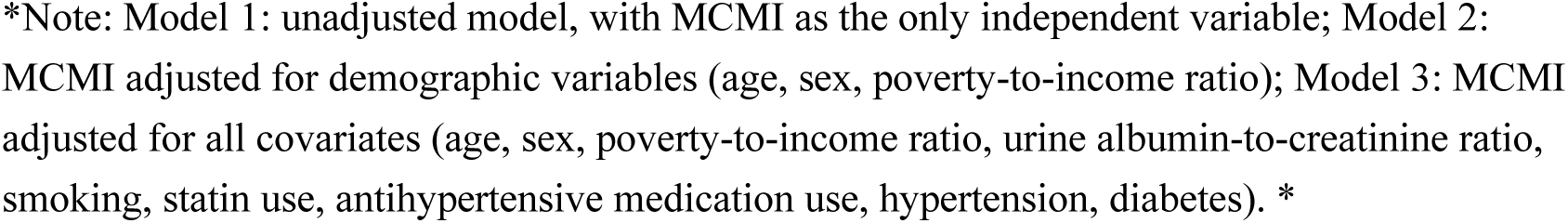
Cox Regression Analysis of MCMI with All-Cause and Cardiovascular Mortality in the CKM Population.

Following full adjustment for all covariates, each one-unit increase in MCMI remained associated with a 21.6% increased risk of all-cause mortality (HR=1.216, 95% CI: 1.070-1.380, p<0.001), but the association with cardiovascular mortality was no longer statistically significant (HR=1.204, 95% CI: 0.948-1.531, p=0.192).

The non-significant association between MCMI and cardiovascular mortality in the fully adjusted model suggested that the effect of MCMI on cardiovascular mortality might not be direct, but rather mediated by metabolism-related confounders such as diabetes, hypertension, and dyslipidemia. Consequently, we repeated the Cox analysis for MCMI and cardiovascular mortality after removing diabetes from the covariates. As shown in Table 8, after excluding diabetes, MCMI demonstrated a significant positive association with cardiovascular mortality risk (HR=1.374, 95% CI: 1.093-1.727, p<0.001). Compared with the model adjusting for diabetes (HR=1.204, p=0.192), these results indicate that diabetes is a key mediator of the association between MCMI and cardiovascular mortality.

**Table 8.**
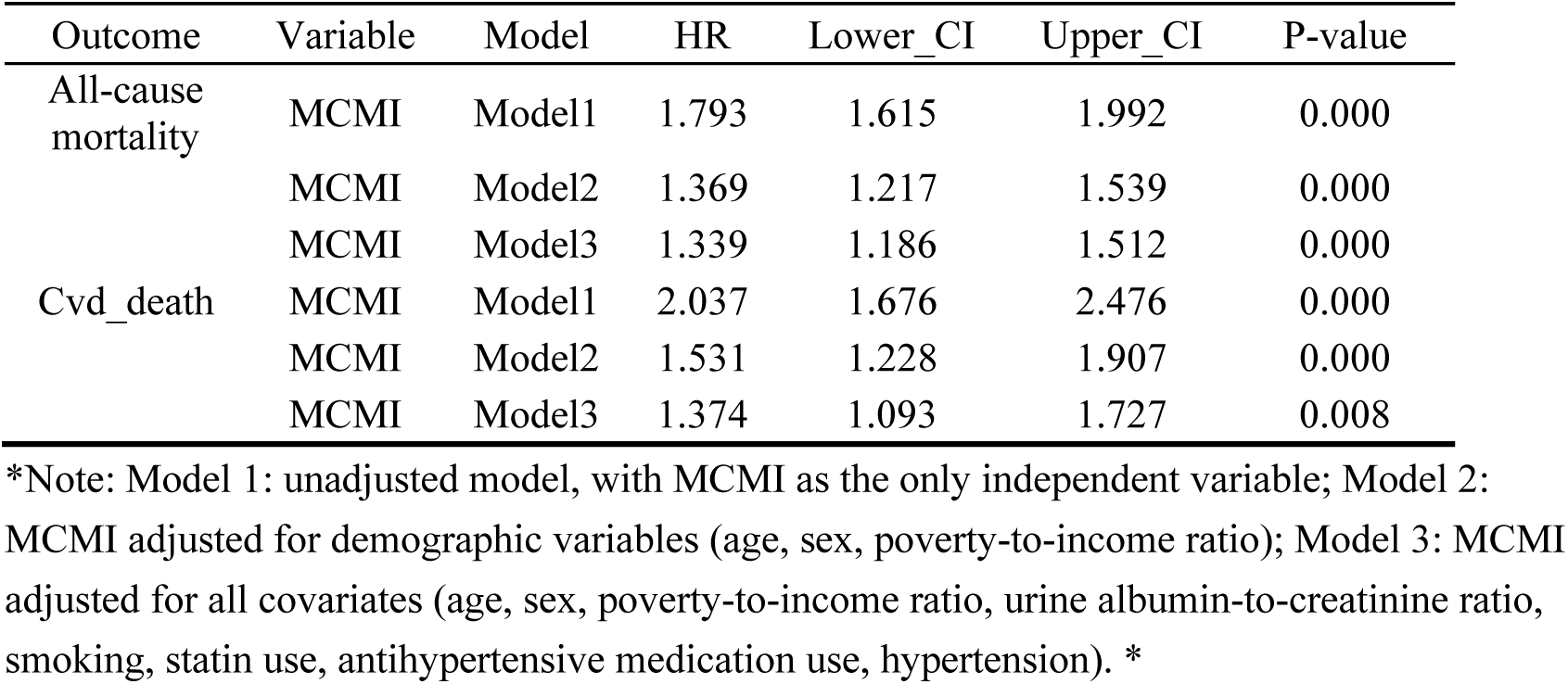
Cox Regression Analysis of MCMI with All-Cause and Cardiovascular Mortality in the CKM Population.

To clarify the mediating role of diabetes in the association between MCMI and cardiovascular mortality, this study employed the gold-standard mediation analysis method based on Cox proportional hazards regression to evaluate the total effect, direct effect, and indirect effect (via diabetes) of MCMI on cardiovascular mortality. After adjusting for confounders including age, sex, poverty-to-income ratio (INDFMPIR), urine albumin-to-creatinine ratio (UACR), smoking status, statin use, and antihypertensive medication use, and incorporating sampling weights, the results showed that the total effect of MCMI on cardiovascular mortality was significant (HR=1.374, 95% confidence interval: 1.093-1.727, P=0.008). After further including diabetes as a mediator, the direct effect of MCMI on cardiovascular mortality was attenuated and no longer statistically significant (HR=1.204, 95% confidence interval: 0.948-1.531, P=0.192).

Quantification using classical epidemiological formulas revealed that the indirect effect of MCMI on cardiovascular mortality mediated by diabetes was HR=1.141, ultimately confirming that diabetes mediated 45.5% of the effect of MCMI on cardiovascular mortality risk (Figure 3).

**Figure 3.**
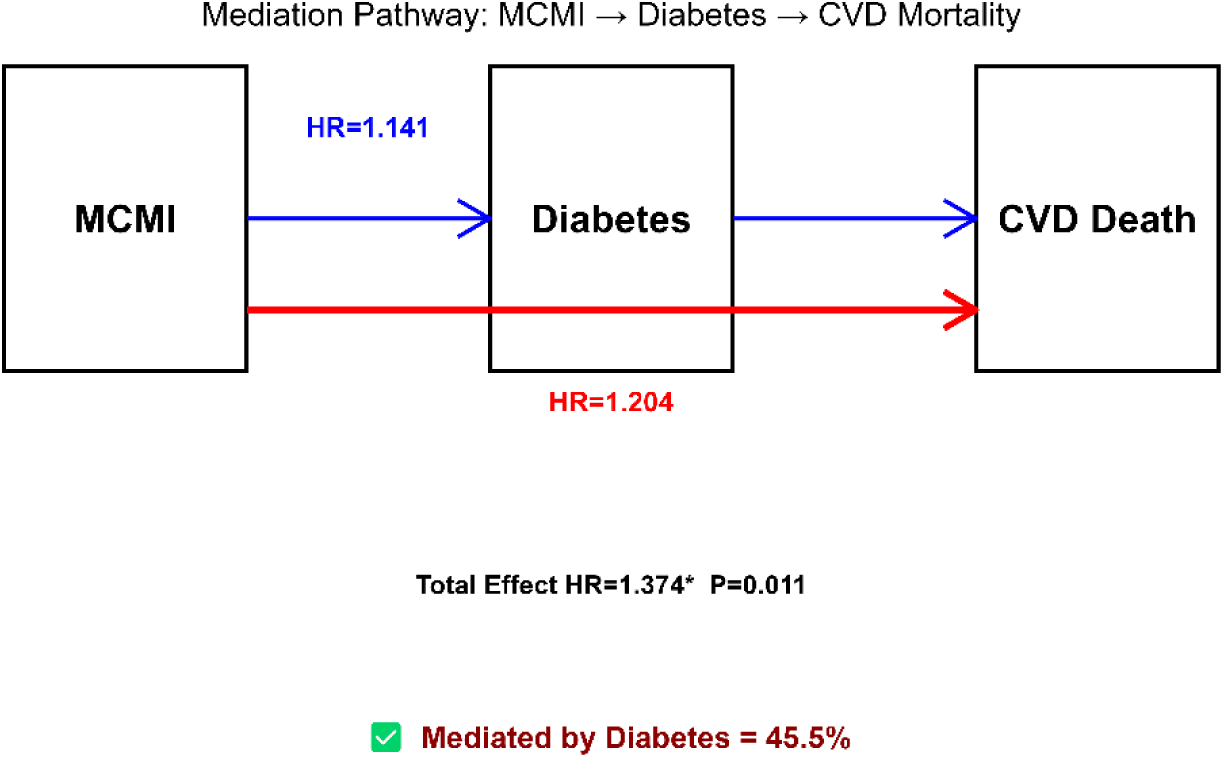
Mediation analysis of diabetes in the effect of MCMI on cardiovascular mortality in the CKM population *Note: The blue line segment in the figure represents the indirect effect pathway (MCMI → diabetes → cardiovascular death), with HR=1.141 indicating the indirect effect risk ratio of MCMI mediated by diabetes. The bold red line segment represents the direct effect pathway (MCMI directly acting on cardiovascular death), with HR=1.204 indicating the direct effect risk ratio after excluding the mediation of diabetes. The boxes represent the core variable nodes, which are, in order, MCMI (modified cardiometabolic index), Diabetes (mediator variable), and CVD Death (outcome variable). The total effect (HR=1.374, P=0.011) represents the overall risk effect of MCMI on cardiovascular mortality, and the mediation proportion of 45.5% represents the quantified mediating effect of diabetes in this association, indicating that 45.5% of the increased cardiovascular mortality risk attributable to elevated MCMI is mediated through the diabetes pathway. *

#### 3.3.2 Association Between MCMI Quartiles and Mortality in the CKM Population

As shown in Table 9, after full adjustment for all confounders (Model 3), compared with the Q1 group, participants in the highest MCMI quartile (Q4) had a significantly increased risk of all-cause mortality (HR=1.412, 95% CI: 1.046-1.907, P=0.024), while no significant differences were observed for the Q2 and Q3 groups. For cardiovascular mortality, none of the MCMI quartile groups showed statistically significant hazard ratios in the fully adjusted model (all P>0.05, with 95% CIs encompassing 1). This contradicted the earlier continuous variable Cox regression analysis in the CKM population, suggesting a non-linear relationship between MCMI and both all-cause and cardiovascular mortality in this population.

**Table 9.**
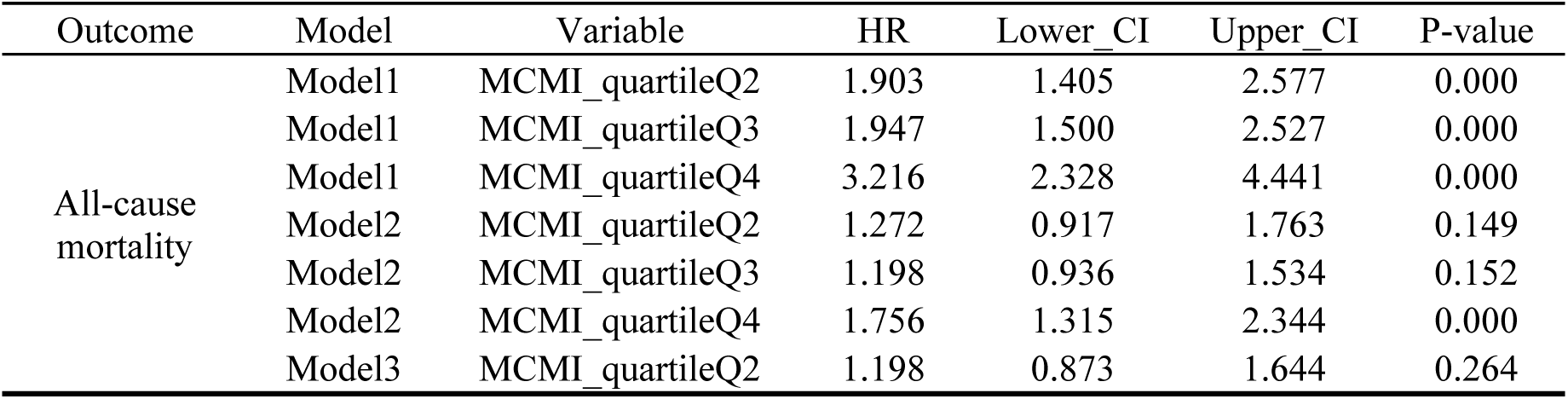
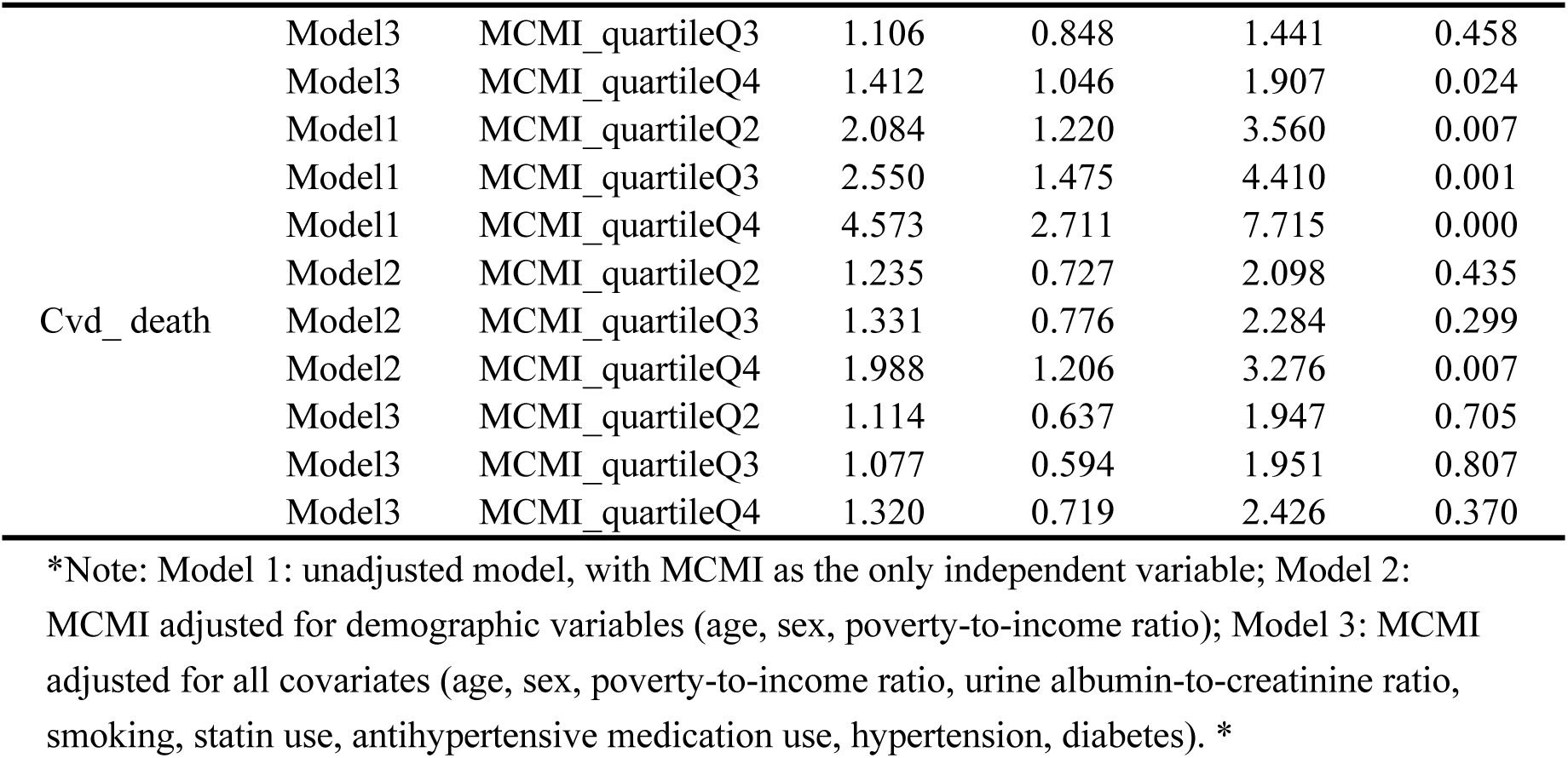
Cox Regression Analysis of MCMI Quartiles with Mortality in the CKM Population.

#### 3.3.3 Testing of Cox Regression Models for MCMI and Mortality in the CKM Population: Schoenfeld Residuals Analysis

We used Schoenfeld residual analysis to test the proportional hazards assumption for the Cox proportional hazards models examining the associations of MCMI with all-cause and cardiovascular mortality in the CKM population. As shown in Figures 4 to 7, the Schoenfeld residuals for MCMI and all covariates in Model 3 were randomly and uniformly distributed around the y=0 axis, and the time trend lines for the residuals (red fitted lines) were approximately horizontal, with all P-values > 0.05. This indicates that the residuals did not change over time, suggesting that the Cox models constructed in this study satisfied the proportional hazards assumption and that the model results are reliable.

**Figure 4.**
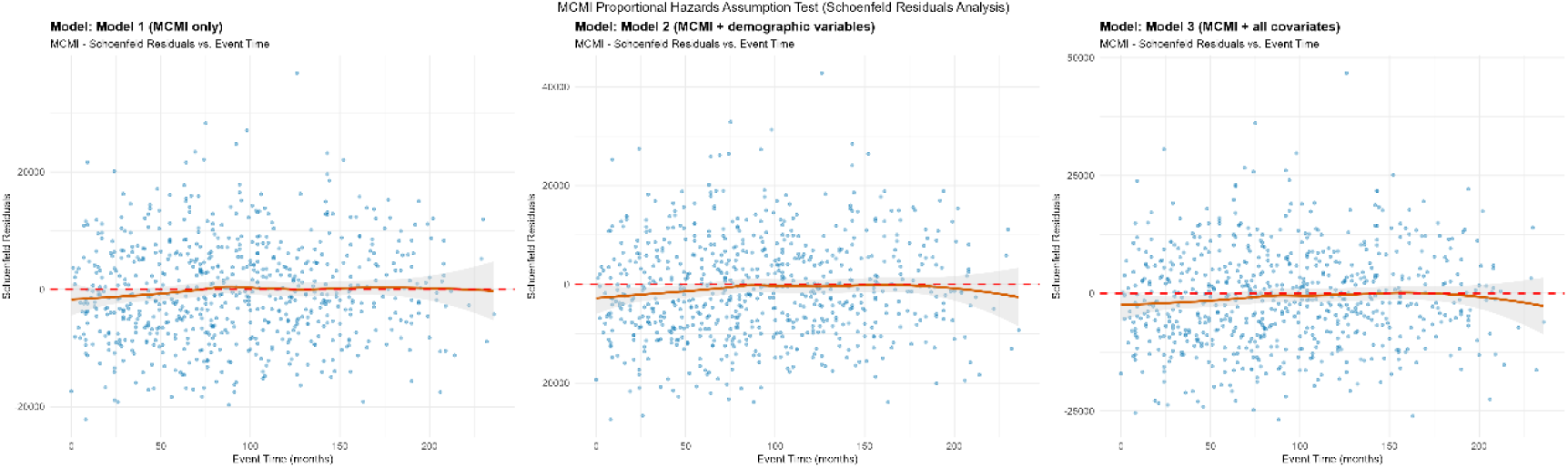
Schoenfeld residuals analysis plot for the Cox proportional hazards model of MCMI with all-cause mortality in the CKM population *Note: This figure shows the distribution of Schoenfeld residuals for MCMI over event time (months) in three progressively adjusted Cox models. Scatter points represent residual values at each event occurrence, and the red fitted line represents the time trend line of the residuals. The three models are: Model 1 (MCMI only), Model 2 (MCMI + demographic variables), and Model 3 (MCMI + all covariates). *

**Figure 5.**
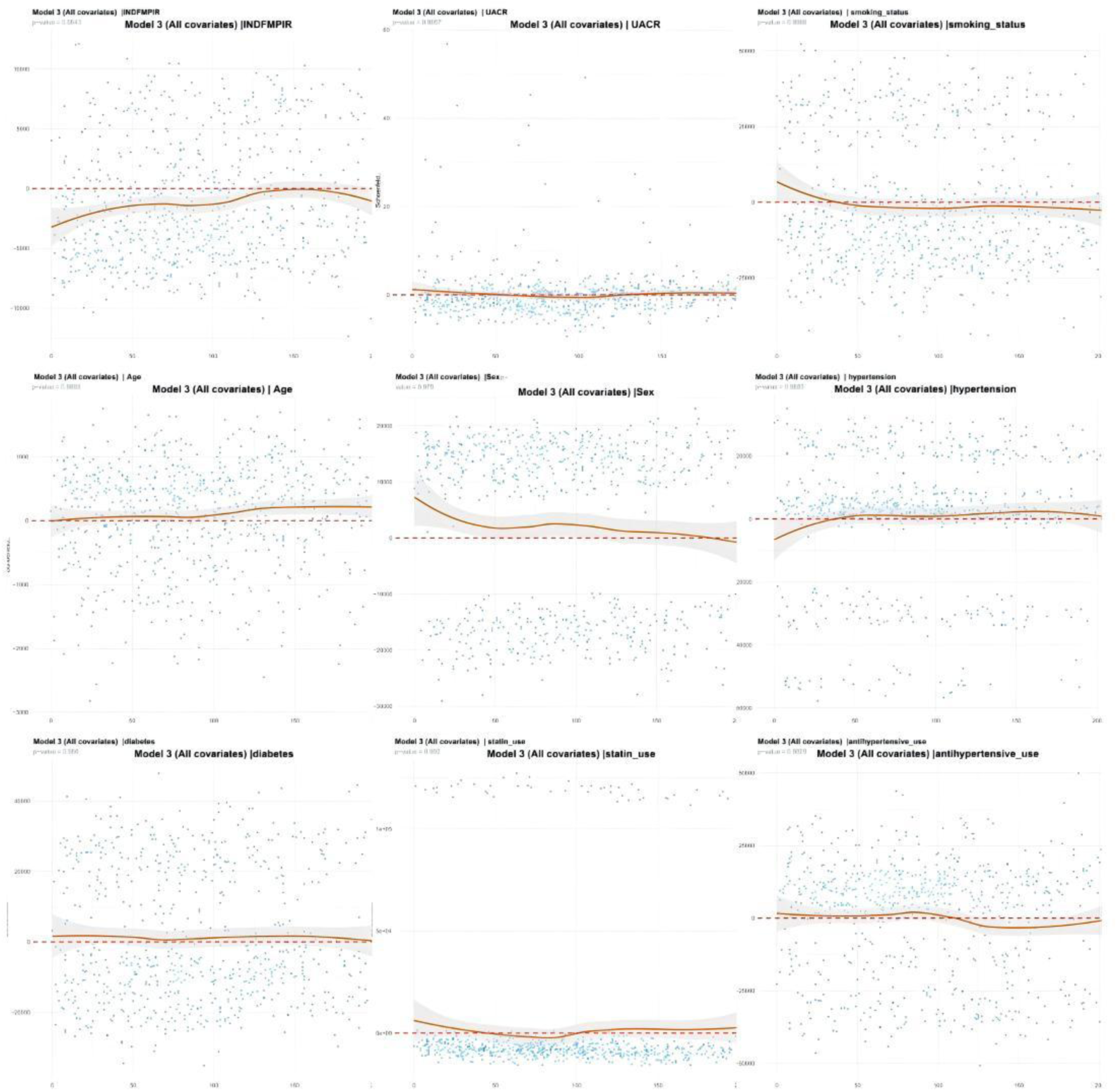
Schoenfeld residuals analysis for all covariates in Model 3 of all-cause mortality Cox proportional hazards model. Note: Scatter points represent residual values at each event occurrence, and the red fitted line represents the time trend line of the residuals.

**Figure 6.**
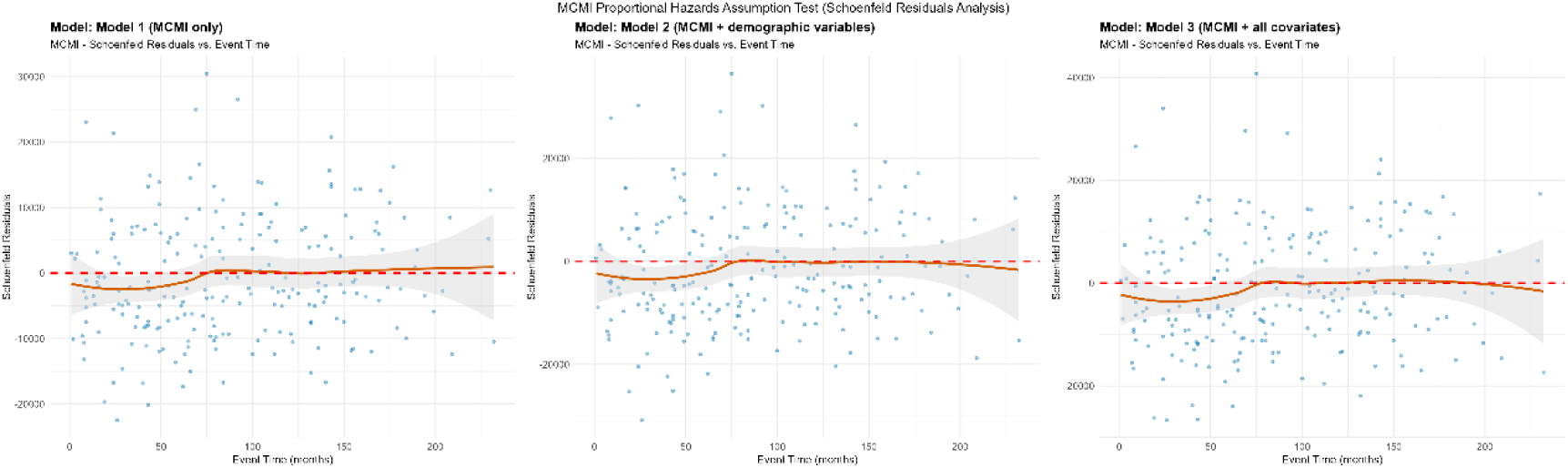
Schoenfeld residuals analysis plot for the Cox proportional hazards model of MCMI with cardiovascular mortality in the CKM population. Note: This figure shows the distribution of Schoenfeld residuals for MCMI over event time (months) in three progressively adjusted Cox models. Scatter points represent residual values at each event occurrence, and the red fitted line represents the time trend line of the residuals. The three models are: Model 1 (MCMI only), Model 2 (MCMI + demographic variables), and Model 3 (MCMI + all covariates).

**Figure 7.**
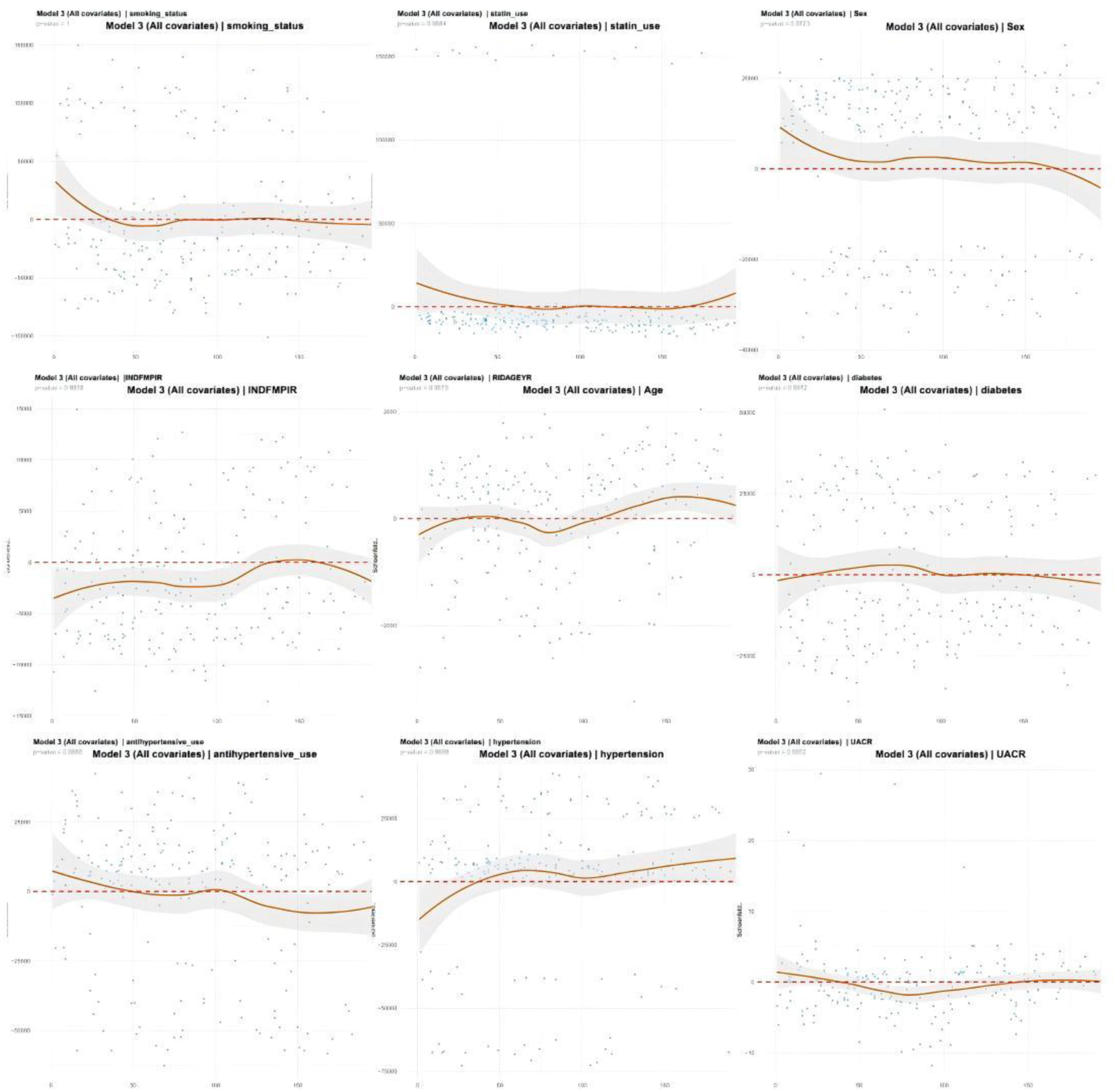
Schoenfeld residuals analysis plot for all covariates in Model 3 of cardiovascular mortality Cox proportional hazards model. Note: Scatter points represent residual values at each event occurrence, and the red fitted line represents the time trend line of the residuals.

### 3.4 Non-Linear Association Between MCMI and All-Cause and Cardiovascular Mortality in the CKM Population

We further employed restricted cubic spline (RCS) analysis to examine the relationship between MCMI and mortality. Figure 8 presents the dose-response relationship curves between the modified cardiometabolic index and mortality in the CKM population (with models adjusted for all covariates), with three knots selected based on the Akaike information criterion (AIC). As shown in Figure 8-A, all-cause mortality risk in the CKM population was positively associated with MCMI, demonstrating a highly significant non-linear relationship. For all-cause mortality, the RCS curve exhibited a monotonically increasing non-linear trend, with a clearly defined safety range (HR < 1 when MCMI < 3.5). When MCMI exceeded 3.5, the HR value continued to increase and remained above 1, confirming that MCMI is an independent risk factor for all-cause mortality with interval-specific risk effects, consistent with the results of the previous Cox regression analyses.

**Figure 8.**
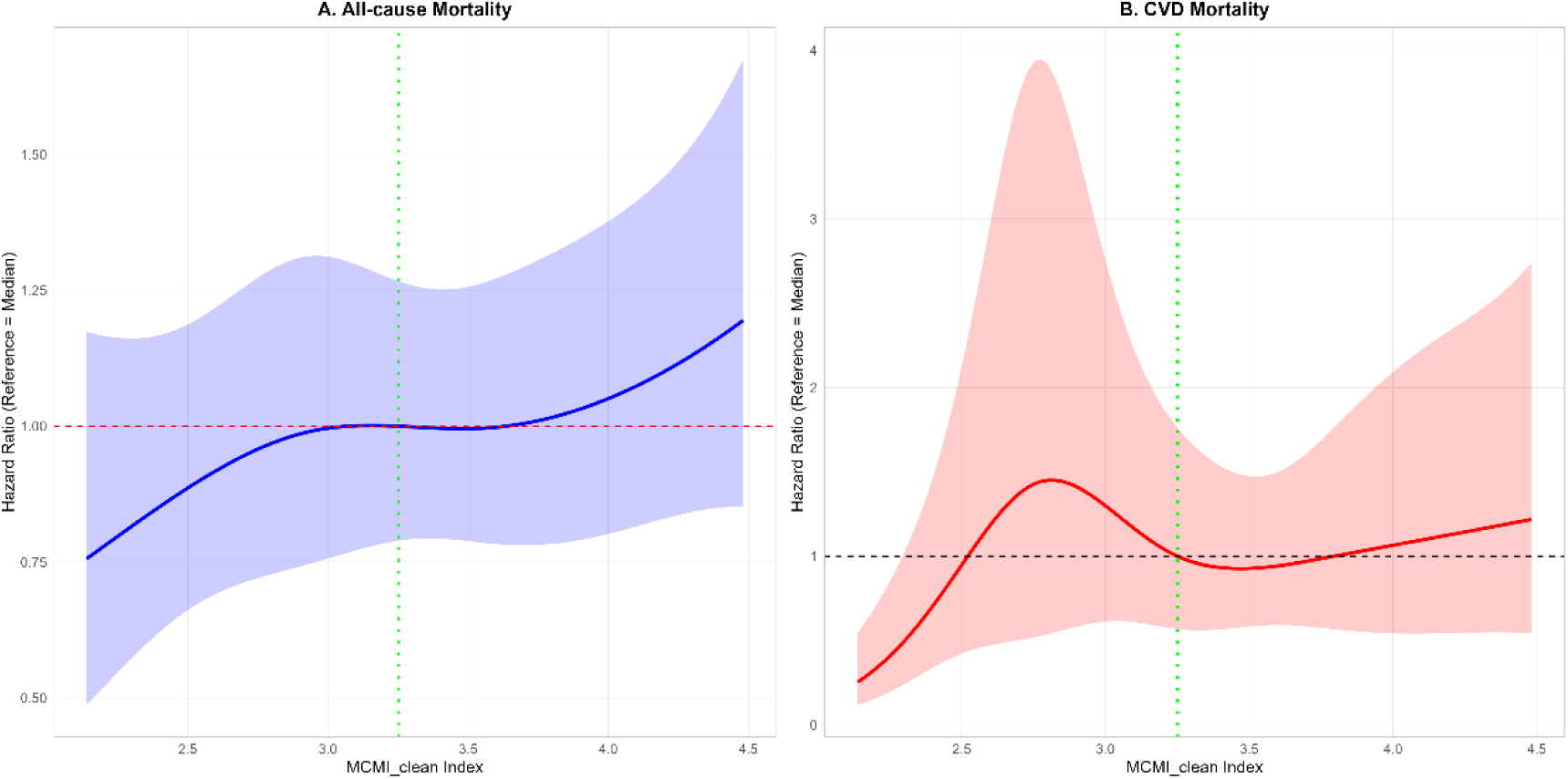
Restricted cubic spline curves for the association between MCMI and all-cause and cardiovascular mortality risk in the CKM population (adjusted for all covariates, including diabetes) *Note: Restricted cubic spline (RCS) models were used to evaluate the non-linear association between the continuous variable MCMI and mortality risk. Models were adjusted for covariates including age, sex, and income, with all variables having a variance inflation factor (VIF) < 5 to exclude severe multicollinearity interference. The curves are referenced to the median MCMI value (HR=1, black dashed line), and the shaded areas represent 95% confidence intervals. Panel A (all-cause mortality): The RCS curve shows a monotonically increasing trend, with HR<1 when MCMI<3.5 (safety range) and HR continuously increasing when MCMI>3.5. Panel B (cardiovascular mortality): The curve shows a non-linear positive association, with HR<1 in the MCMI 3.0–3.5 range (low-risk interval) and HR>1 when MCMI<2.5 or >4.0 (high-risk intervals).*

Figure 8-B shows the RCS analysis curve for the association between MCMI and cardiovascular mortality risk in the CKM population (with four knots selected based on the AIC criterion). The curve exhibited a “rise-fall-rise” pattern, with the overall trend remaining positively correlated. This curve also identified low-risk (MCMI 3.0-3.5) and high-risk (MCMI < 2.5 or > 4.0) intervals, indicating that the risk effect of MCMI on cardiovascular mortality also has significant interval-specific characteristics. Although the Cox regression analysis of MCMI quartiles with cardiovascular mortality in the CKM population did not show significant associations, this result essentially reflects the loss of continuous non-linear information due to categorical variable grouping. Figure 9 presents the RCS curves after adjusting for all covariates except diabetes. Compared with Figure 1-8, the curve trends in Figure 9 were identical, but the risk effect of MCMI on mortality outcomes in the CKM population was more pronounced. Combined with the aforementioned mediation analysis of MCMI and cardiovascular mortality (diabetes mediating 45.5% of the risk effect), these findings confirm that MCMI remains an independent risk factor for cardiovascular mortality, with diabetes serving as an important mediating pathway for the MCMI risk effect.

**Figure 9.**
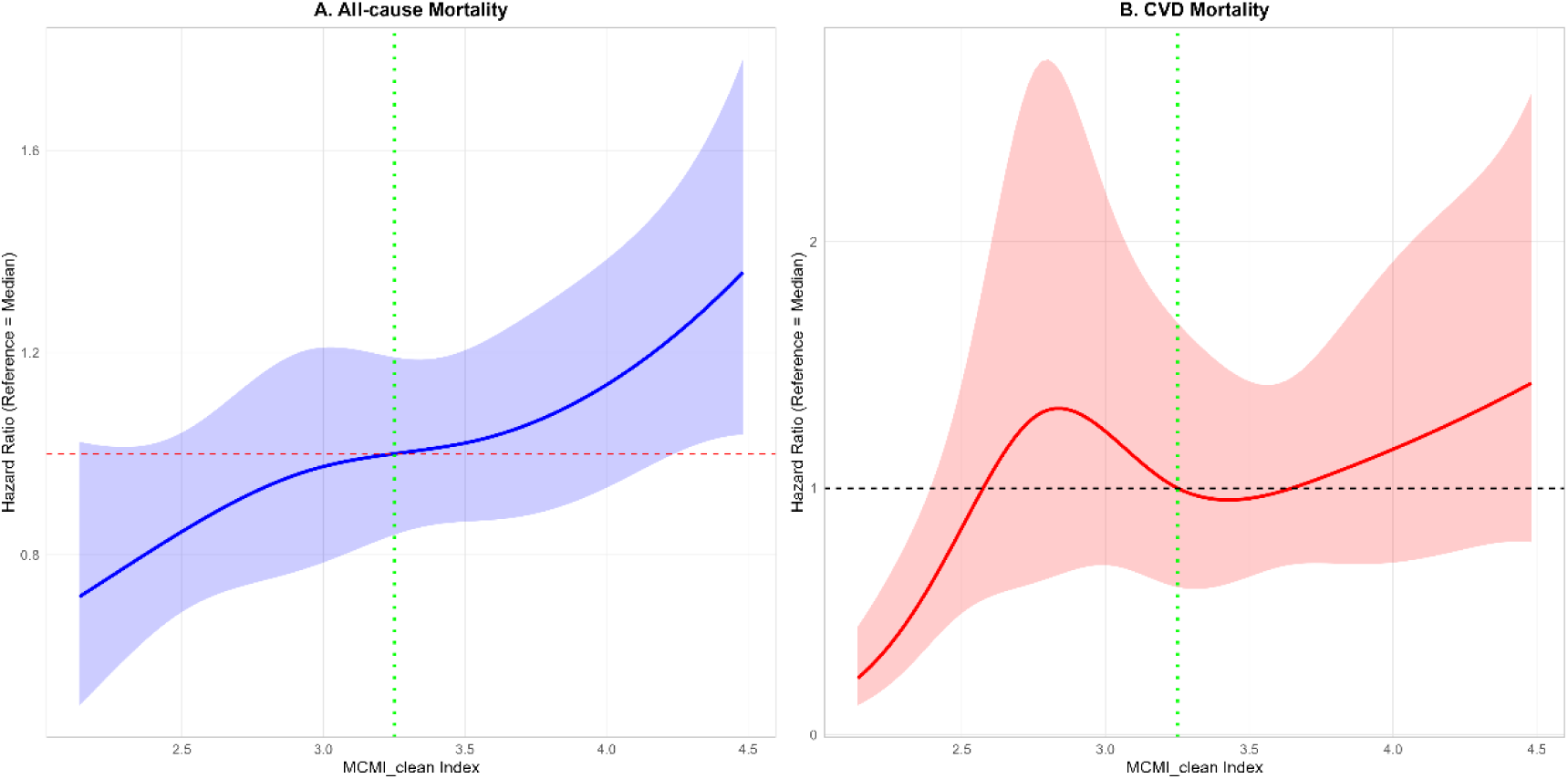
Restricted cubic spline curves for the association between MCMI and all-cause and cardiovascular mortality risk in the CKM population (adjusted for all covariates, excluding diabetes) *Note: Model specifications are identical to those in Figure 1, with the only difference being the exclusion of the diabetes covariate. The RCS curve trends for Panel A (all-cause mortality) and Panel B (cardiovascular mortality) are highly consistent with those in Figure 1, suggesting that the independent effect of MCMI on mortality risk is not entirely confounded by diabetes. Compared with quartile-based grouping analysis, the RCS analysis of MCMI as a continuous variable more completely preserves the risk interval characteristics of MCMI, avoiding information loss associated with categorical variable transformation.*

### 3.5 Subgroup Analyses and Interaction Analyses

Figure 10 presents a forest plot of subgroup analyses for the association between the modified cardiometabolic index and all-cause mortality in the CKM population. Subgroup analyses were conducted based on age, sex, race/ethnicity, alcohol consumption, disease status (hypertension, diabetes), and CKM stage (after covariate adjustment). Among individuals aged 45 years and older, the association between MCMI and all-cause mortality was more pronounced (ages 45-59: HR=1.741, 95% CI: 1.262-2.400, P=0.001; ages 60-74: HR=1.269, 95% CI: 1.050-1.533, P=0.014), suggesting that the impact of MCMI on mortality risk may be more prominent in middle-aged and older adults, possibly due to decreased physiological reserve. Both males (HR=1.438, P<0.001) and females (HR=1.228, P=0.023) showed increased mortality risk associated with MCMI, but notable racial/ethnic differences were observed: White individuals (HR=1.303, P=0.001) and Other Hispanic individuals (HR=2.235, P<0.001) demonstrated significantly elevated mortality risk, which may be related to race-specific health disparities such as differences in underlying disease distribution and healthcare access. Among individuals with hypertension (HR=1.31, 95% CI: 1.13-1.51, P<0.001), the association between MCMI and mortality risk was more pronounced, suggesting that hypertension may be a synergistic factor amplifying the mortality risk associated with MCMI. The association between MCMI and all-cause mortality exhibited population heterogeneity, with higher risks observed in older adults, specific racial/ethnic groups, alcohol consumers, individuals with hypertension not using antihypertensive medication, those not using statins, and those with advanced CKM stages.

**Figure 10.**
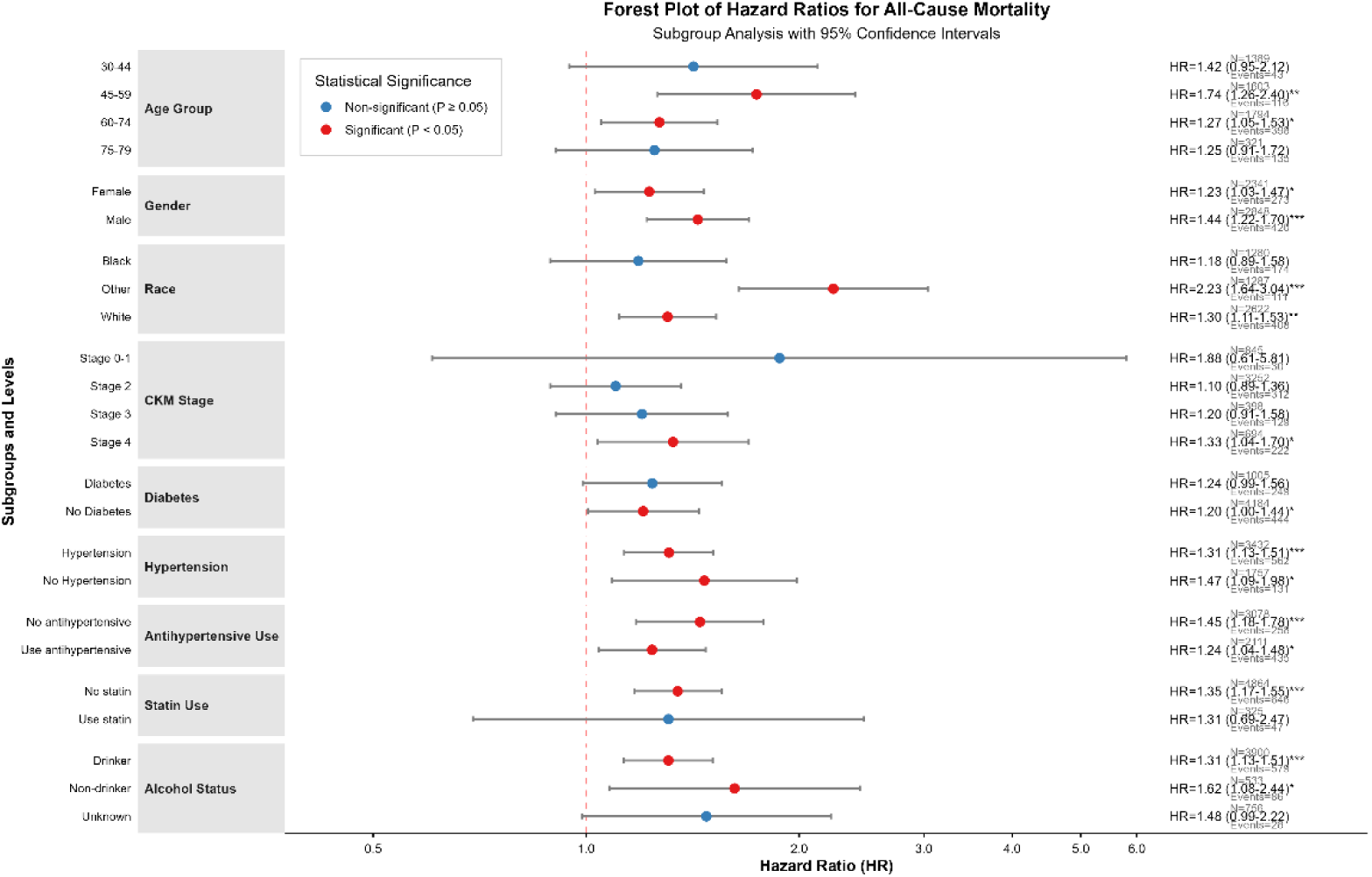
Forest plot of subgroup analyses for the association between MCMI and all-cause mortality risk in the CKM population *Note: After subgroup analyses stratified by age, sex, race/ethnicity, alcohol consumption, disease status, and CKM stage (with covariate adjustment), the association between MCMI and all-cause mortality exhibited population heterogeneity: higher mortality risks were observed in middle-aged and older adults, specific racial/ethnic groups, alcohol consumers, individuals with hypertension not using antihypertensive medication, those not using statins, and those with advanced CKM stages. The associations were particularly pronounced in individuals aged >45 years, those with hypertension, and certain Hispanic subgroups.*

Figure 11 displays a forest plot of subgroup analyses for the association between the modified cardiometabolic index and cardiovascular disease (CVD) mortality in the CKM population. As shown in the figure, the association between MCMI and CVD mortality also exhibited population heterogeneity, with the strength of the association influenced by sex, race/ethnicity, alcohol consumption status, and comorbid conditions. Notably, male sex, alcohol consumption, hypertension, hypertension without antihypertensive medication use, and non-use of statins all synergistically increased the cardiovascular mortality risk associated with MCMI in the CKM population. The lack of statistically significant associations in some subgroups was primarily attributable to insufficient statistical power due to a small number of events, rather than indicating an absence of risk effects for MCMI.

**Figure 11.**
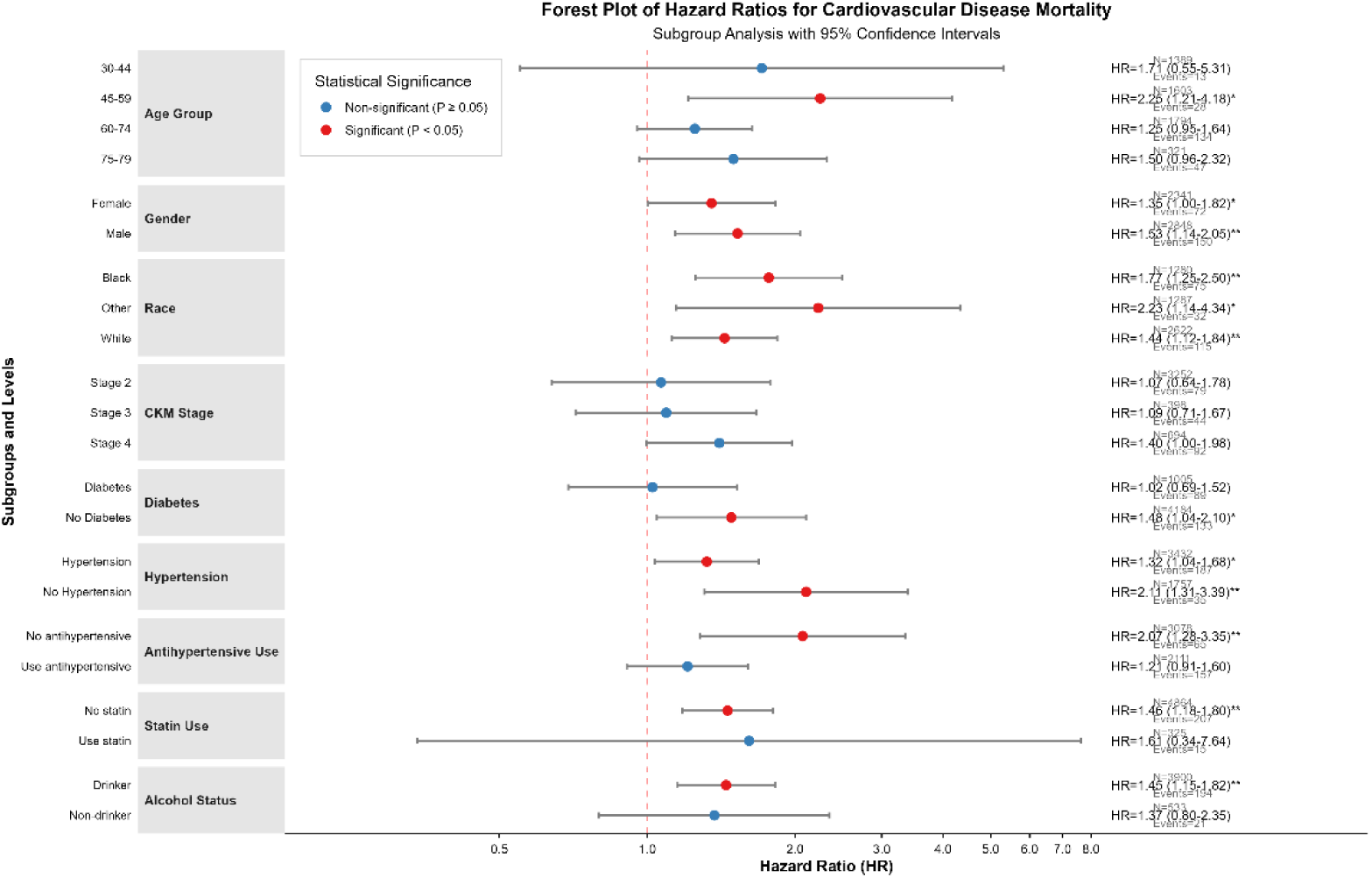
Forest plot of subgroup analyses for the association between MCMI and cardiovascular mortality risk in the CKM population *Note: The association between MCMI and CVD mortality also exhibited population heterogeneity, with the strength of the association influenced by sex, race/ethnicity, alcohol consumption status, and comorbid conditions. Male sex, alcohol consumption, hypertension, hypertension without antihypertensive medication use, and non-use of statins all synergistically increased the cardiovascular mortality risk associated with MCMI in the CKM population. The lack of statistically significant associations in some subgroups was primarily attributable to insufficient statistical power due to a small number of events, rather than indicating an absence of risk effects for MCMI. *

To further evaluate subgroup heterogeneity in the association between MCMI and mortality risk, this study tested potential effect modification by including interaction terms. Figure 12 presents an interaction heatmap, from which it can be clearly observed that in the all-cause mortality model, all interaction terms—including age subgroup, alcohol consumption status subgroup, and CKM stage subgroup—had P-values > 0.05, indicating no significant heterogeneity in the effect of MCMI on all-cause mortality across subgroups and confirming the robustness of MCMI as an independent risk factor for all-cause mortality. In the cardiovascular mortality model, only the interaction term for the CKM stage subgroup showed statistical significance, suggesting that CKM stage significantly modifies the cardiovascular risk associated with MCMI. Overall, the association between MCMI and mortality risk in the CKM population showed no significant heterogeneity across subgroups, further confirming the robustness of MCMI as an independent risk factor.

**Figure 12.**
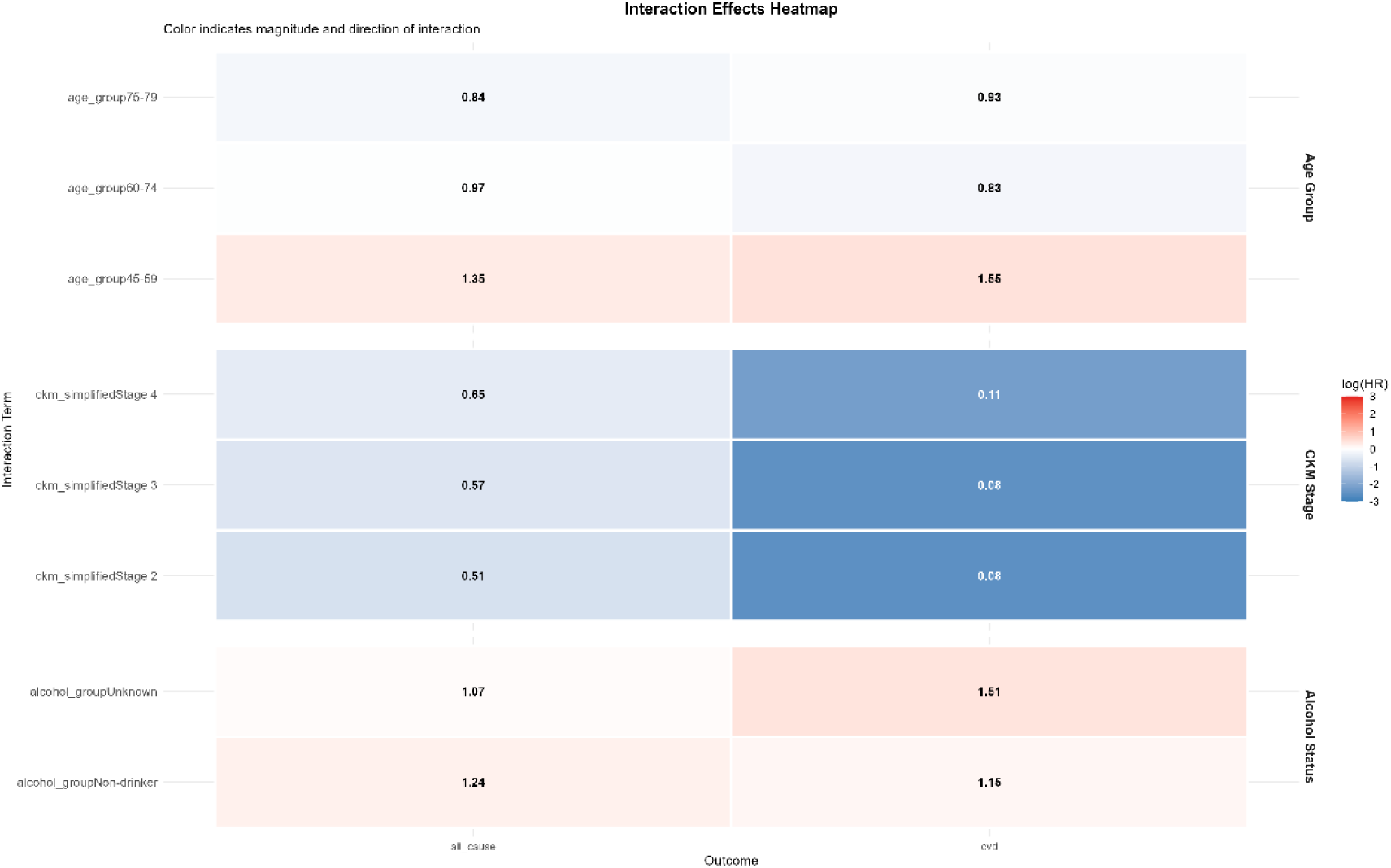
Subgroup interaction heatmap for the association between MCMI and mortality risk in the CKM population *Note: The values in the figure represent P-values, with P < 0.05 indicating statistical significance. Different colors in the heatmap represent log(HR), i.e., the log-transformed hazard ratio values. Overall, the effect of MCMI on mortality risk in the CKM population showed no significant heterogeneity across subgroups.*

### 3.6 Sensitivity Analyses

#### 3.6.1 Analysis by Different Categorical Classifications of MCMI

To verify that MCMI is an independent risk factor in the CKM population, we further performed Cox regression analyses by applying different grouping strategies for MCMI in relation to all-cause mortality risk. As shown in Table 10, continuous variable analysis revealed that each one-unit increase in MCMI was associated with a 22% increased risk of mortality (HR=1.22, 95% CI: 1.05-1.40, p=0.008). When using quartile grouping, we observed a clear dose-response trend, with only the highest quartile (Q4) showing a significantly increased mortality risk of 43% (HR=1.43, 95% CI: 1.06-1.93, p=0.020), while Q2 and Q3 showed no statistically significant differences compared to Q1 (both p>0.05). To further validate this “threshold effect,” we conducted quintile grouping analysis, which further confirmed that only the highest quintile (Q5) had a significantly increased mortality risk of 55% (HR=1.55, 95% CI: 1.12-2.16, p=0.009), whereas tertile grouping failed to detect significant associations due to overly coarse categorization. Analysis using a simple median dichotomization also revealed no significant association (HR=0.88, p=0.242), suggesting that the risk effect of MCMI is primarily concentrated in the higher range of its distribution.

**Table 10.** Cox Regression Analysis of Differently Grouped MCMI with All-Cause Mortality in the CKM Population.

| Grouping | Comparison | HR | Lower_CI | Upper_CI | P-Value |
| --- | --- | --- | --- | --- | --- |
| Continuous | MCMI | 1.216 | 1.052 | 1.404 | 0.008 |
| Quartile | Q2 vs Q1 | 1.202 | 0.889 | 1.624 | 0.231 |
| Quartile | Q3 vs Q1 | 1.136 | 0.865 | 1.492 | 0.361 |
| Quartile | Q4 vs Q1 | 1.429 | 1.059 | 1.930 | 0.020 |
| Tertile | T2 vs T1 | 1.065 | 0.820 | 1.384 | 0.637 |
| Tertile | T3 vs T1 | 1.201 | 0.915 | 1.576 | 0.187 |
| Quintile | Q2 vs Q1 | 1.331 | 0.943 | 1.880 | 0.104 |
| Quintile | Q3 vs Q1 | 1.189 | 0.859 | 1.646 | 0.297 |
| Quintile | Q4 vs Q1 | 1.150 | 0.848 | 1.560 | 0.368 |
| Quintile | Q5 vs Q1 | 1.552 | 1.117 | 2.156 | 0.009 |
| Binary | High vs Low | 0.878 | 0.706 | 1.092 | 0.242 |

\*Note: Continuous represents MCMI analyzed as a continuous variable; Quartile represents MCMI grouped into quartiles; Tertile represents MCMI grouped into tertiles; Quintile represents MCMI grouped into quintiles; Binary represents MCMI dichotomized into low and high groups based on the median value. The model was adjusted for the same covariates as Model 3 described previously (adjusted for age, sex, poverty-to-income ratio, urine albumin-to-creatinine ratio, smoking, statin use, antihypertensive medication use, hypertension, and diabetes). \*

#### 3.6.2 Comparative Analysis After Excluding Specific Populations

To exclude potential bias, we also performed a series of sensitivity analyses by exclusion. After excluding individuals with follow-up less than 1 year (n=5,153), those with baseline cancer diagnosis (n=4,611), or those with severe renal insufficiency (eGFR <30 mL/min/1.73m²) (n=5,176), the positive association between MCMI and all-cause mortality risk remained stable, with HRs ranging from 1.22 to 1.23 and all P-values <0.05. Even when the analysis was restricted to non-Hispanic White individuals (n=2,622), the results remained consistent (HR=1.20, 95% CI: 1.02-1.42, p=0.026), demonstrating the robustness of the findings across populations with different characteristics (see Table 11 for details).

**Table 11.** Cox Regression Analysis of MCMI with All-Cause Mortality in the CKM Population After Excluding Specific Populations.

| Analysis | HR | Lower_CI | Upper_CI | P_Value | N |
| --- | --- | --- | --- | --- | --- |
| Exclude Followup <1year | 1.221 | 1.053 | 1.415 | 0.008 | 5153 |
| Exclude Baseline Cancer | 1.222 | 1.037 | 1.441 | 0.017 | 4611 |
| Exclude eGFR < 30 | 1.223 | 1.058 | 1.413 | 0.006 | 5176 |
| White Only | 1.203 | 1.022 | 1.416 | 0.026 | 2622 |

\*Note: Exclude Followup <1 year represents the exclusion of individuals with follow-up less than 1 year (n=5,153); Exclude Baseline Cancer represents the exclusion of individuals with baseline cancer diagnosis (n=4,611); Exclude eGFR <30 represents the exclusion of individuals with severe renal insufficiency (eGFR <30 mL/min/1.73m<sup>2</sup>) (n=5,176); White Only represents analysis restricted to non-Hispanic White individuals (n=2,622). The model was adjusted for the same covariates as Model 3 described previously (adjusted for age, sex, poverty-to-income ratio, urine albumin-to-creatinine ratio, smoking, statin use, antihypertensive medication use, hypertension, and diabetes).\*

#### 3.6.3 Competing Risk Model Analysis

Given the competing risk between cardiovascular disease (CVD) and non-CVD mortality, we further employed the Fine-Gray proportional subdistribution hazards model, focusing on CVD mortality outcomes. The results showed that after accounting for competing risks, the strength of the association between MCMI and CVD mortality increased substantially, demonstrating a clear dose-response relationship (Table 12): compared with the Q1 group, the subdistribution hazard ratios (sHRs) for the Q2, Q3, and Q4 groups were 1.88, 2.47, and 3.25, respectively (trend P < 0.001). The cumulative incidence function curves (Figure 13) visually demonstrated a gradient increase in the cumulative incidence of CVD mortality with ascending MCMI quartiles. In contrast, results from cause-specific Cox models (Table 13) showed that the associations between MCMI and CVD mortality did not reach statistical significance (all P > 0.05). This indicates that failure to account for competing non-CVD mortality events may substantially underestimate the true risk effect of MCMI on CVD mortality. In conclusion, MCMI is an independent risk factor for cardiovascular mortality in the CKM population.

**Figure 13.**
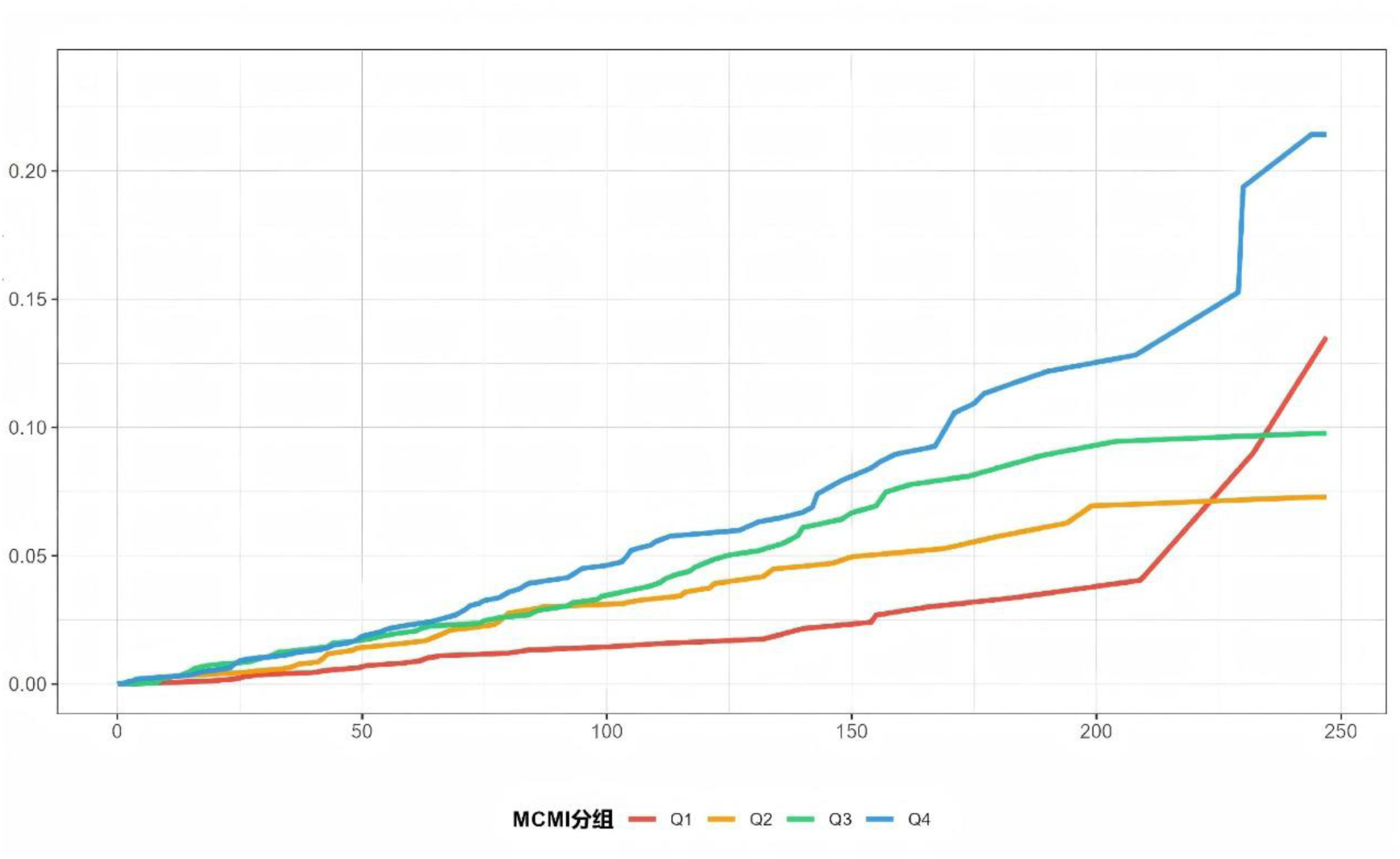
Cumulative incidence curves for cardiovascular mortality risk in the CKM population by MCMI quartiles *Note: The x-axis represents follow-up time (months), and the y-axis represents the cumulative incidence function (CIF) of CVD mortality. It can be intuitively observed from the figure that the cumulative incidence of cardiovascular mortality shows a gradient increasing trend with higher MCMI quartiles and longer follow-up time. *

**Table 12.** Competing risk model analysis of MCMI quartiles with cardiovascular mortality risk in the CKM population.

| Group | Subdistribution HR | Lower 95CI | Upper 95CI | Z-value | P-value |
| --- | --- | --- | --- | --- | --- |
| Q2 vs Q1 | 1.878 | 1.163 | 3.031 | 2.578 | 0.0099 |
| Q3 vs Q1 | 2.466 | 1.563 | 3.889 | 3.882 | 1e-04 |
| Q4 vs Q1 | 3.254 | 2.1 | 5.042 | 5.28 | 0 |

\*Note: It can be clearly observed from the table that the hazard ratios increased substantially with increasing MCMI, indicating a significantly enhanced association between MCMI and CVD mortality. The model was adjusted for the same covariates as Model 3 described previously (adjusted for age, sex, poverty-to-income ratio, urine albumin-to-creatinine ratio, smoking, statin use, antihypertensive medication use, hypertension, and diabetes).\*

**Table 13.** Cause-specific Cox regression model analysis of MCMI quartiles with cardiovascular mortality risk in the CKM population.

| Outcome | Comparison | HR | P-Value |
| --- | --- | --- | --- |
| CVD Death | Q2 vs Q1 | 1.114 | 0.705 |
| CVD Death | Q3 vs Q1 | 1.077 | 0.807 |
| CVD Death | Q4 vs Q1 | 1.320 | 0.370 |
| Non-CVD Death | Q2 vs Q1 | 1.240 | 0.269 |
| Non-CVD Death | Q3 vs Q1 | 1.158 | 0.373 |
| Non-CVD Death | Q4 vs Q1 | 1.484 | 0.042 |

\*Note: The model was adjusted for the same covariates as Model 3 described previously (adjusted for age, sex, poverty-to-income ratio, urine albumin-to-creatinine ratio, smoking, statin use, antihypertensive medication use, hypertension, and diabetes).\*

#### 3.6.4 Model Specification Analysis and E-Value Assessment

We also employed a comparative analysis approach using different covariate adjustment strategies. As shown in Table 14, in the minimal model adjusting only for demographic variables (age and sex), the association between MCMI and mortality risk was strongest (HR=1.47, 95% CI: 1.27-1.71). As covariates including socioeconomic status, lifestyle factors, and clinical comorbidities were progressively incorporated into the models, the hazard ratios exhibited a gradient attenuation trend (from 1.40 in the Basic model to 1.22 in the Full model), indicating that these known factors partially explained the association between MCMI and mortality risk.

**Table 14.**
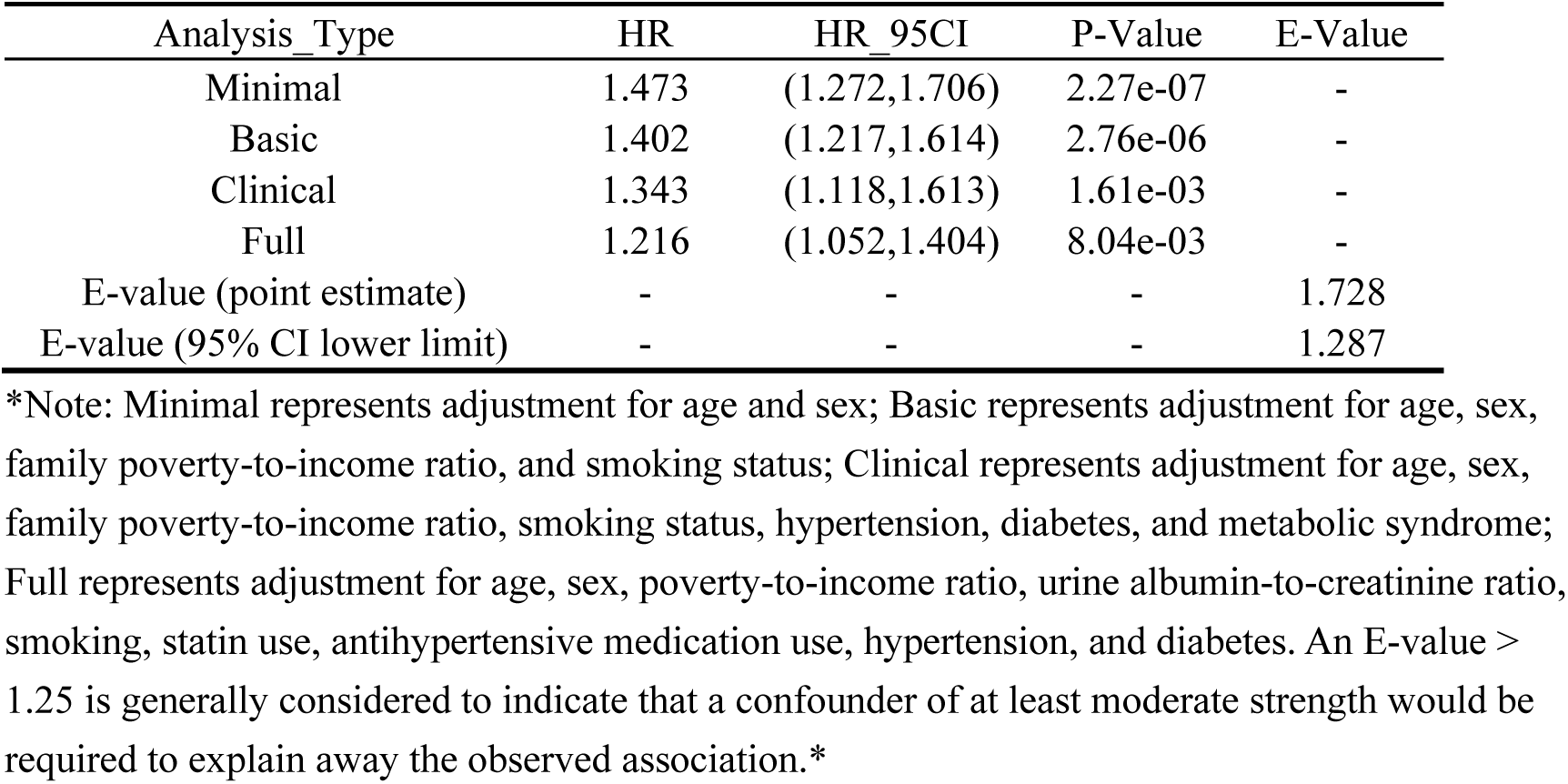
Cox Regression Analysis of MCMI with All-Cause Mortality in the CKM Population Using Different Covariate Adjustment Strategies.

| Analysis_Type | HR | HR_95CI | P-Value | E-Value |
| --- | --- | --- | --- | --- |
| Minimal | 1.473 | (1.272,1.706) | 2.27e-07 | - |
| Basic | 1.402 | (1.217,1.614) | 2.76e-06 | - |
| Clinical | 1.343 | (1.118,1.613) | 1.61e-03 | - |
| Full | 1.216 | (1.052,1.404) | 8.04e-03 | - |
| E-value (point estimate) | - | - | - | 1.728 |
| E-value (95% CI lower limit) | - | - | - | 1.287 |

\*Note: Minimal represents adjustment for age and sex; Basic represents adjustment for age, sex, family poverty-to-income ratio, and smoking status; Clinical represents adjustment for age, sex, family poverty-to-income ratio, smoking status, hypertension, diabetes, and metabolic syndrome; Full represents adjustment for age, sex, poverty-to-income ratio, urine albumin-to-creatinine ratio, smoking, statin use, antihypertensive medication use, hypertension, and diabetes. An E-value > 1.25 is generally considered to indicate that a confounder of at least moderate strength would be required to explain away the observed association.\*

Nevertheless, even in the fully adjusted model with 9 potential confounders, each one-unit increase in MCMI remained independently associated with a 22% increased risk of all-cause mortality (HR=1.22, 95% CI: 1.05-1.40, p=0.008). To assess the potential impact of unmeasured confounding on the study conclusions, we further calculated E-values. The results showed that to fully explain the observed association (HR=1.22), an unmeasured confounder would need to be associated with both MCMI and mortality risk by a hazard ratio of at least 1.73. Even using a conservative estimate of the association strength (the lower limit of the 95% CI, HR=1.05), the required hazard ratio would be no less than 1.29. This further validates the robustness of the association between MCMI and mortality risk, demonstrating considerable robustness against unmeasured confounding.

### 3.7 Comparison of the Independent Predictive Value of MCMI, TyG, and CMI for Prognosis in the CKM Population

To compare the predictive performance of MCMI, TyG, and CMI for mortality risk in the CKM population, we performed Cox regression analyses for each of the three indices individually with all-cause mortality and cardiovascular mortality in the CKM population, and compared their predictive efficacy using the C-index. As shown in Figure 14, MCMI exhibited the optimal predictive performance for both all-cause mortality and cardiovascular mortality. We further analyzed the AUC differences between MCMI and TyG over the 12-year follow-up period (AUC difference = MCMI AUC − TyG AUC) to dynamically evaluate the changes in their predictive advantages. The results showed that the AUC differences were positive at all time points (mean difference = 0.0243), indicating that the predictive accuracy of MCMI consistently outperformed that of TyG throughout the entire follow-up period (Figure 15).

**Figure 14.**
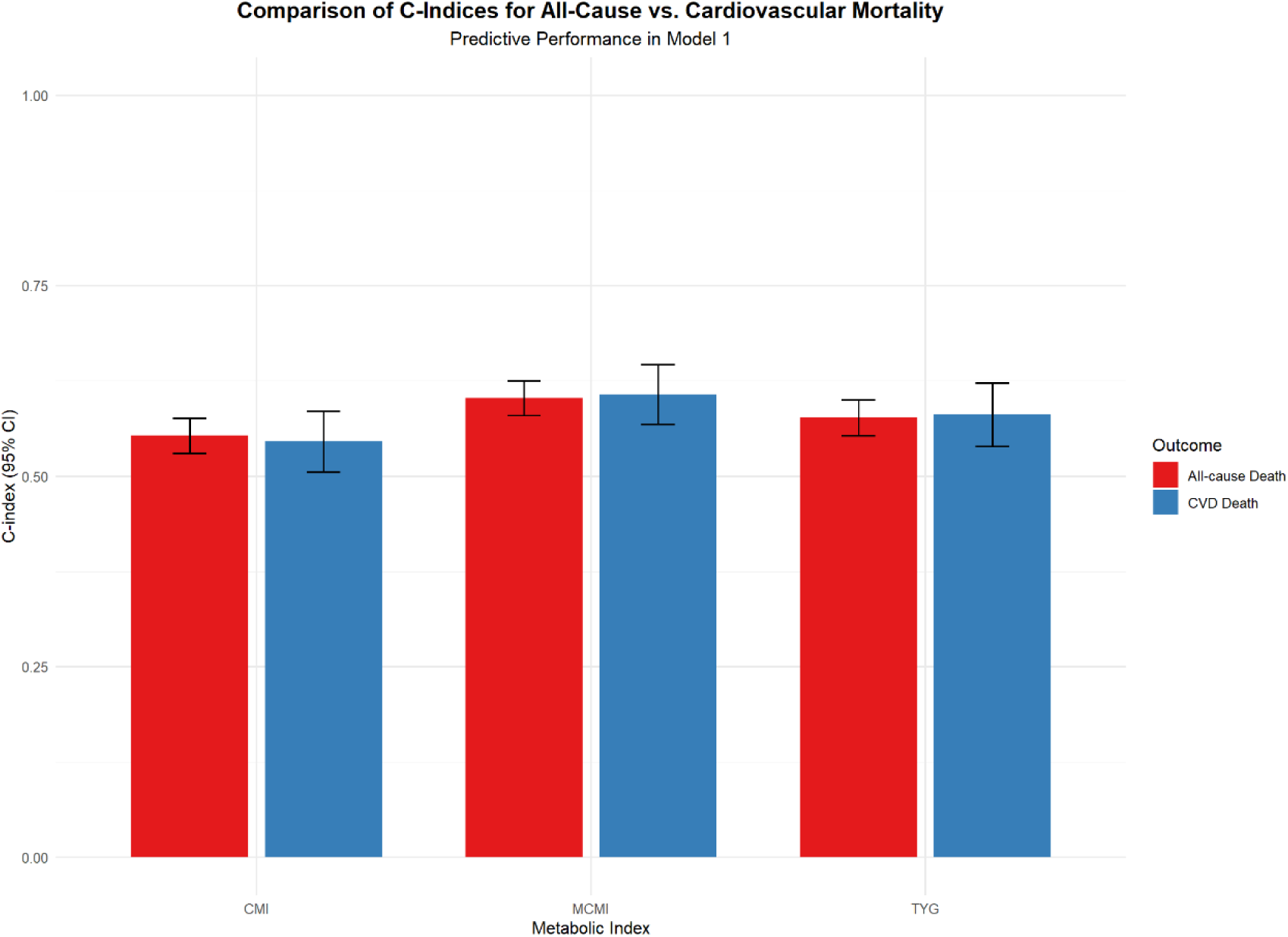
omparison of predictive performance of MCMI, TyG, and CMI for all-cause and cardiovascular mortality risk in the CKM population

**Figure 15.**
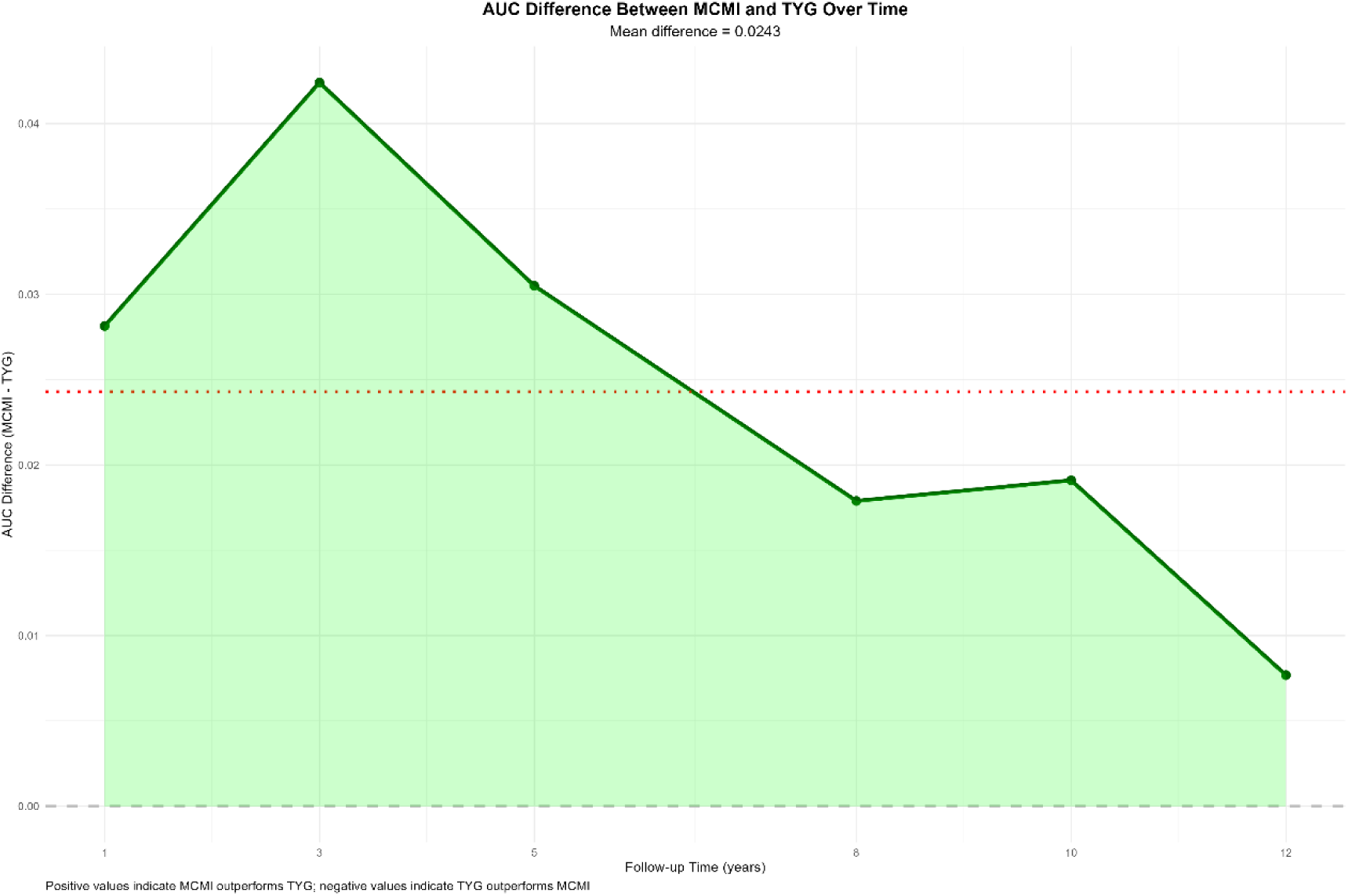
AUC differences between MCMI and TyG over 12 years of follow-up in the CKM population *Note: The y-axis represents the AUC difference (MCMI − TyG), with positive values indicating that the AUC of the MCMI model is higher than that of the TyG model, and negative values indicating the opposite. The red dashed line in the figure represents the mean difference (0.0243), and the green curve shows the trend of AUC differences over 1–12 years of follow-up.*

## 4. Discussion

This study, based on a large-scale community-based cohort of patients with Cardiovascular-Kidney-Metabolic (CKM) syndrome, aimed to elucidate the association of the modified cardiometabolic index (MCMI) with CKM syndrome staging and mortality risk. The study found that MCMI exhibited a “decelerating increase” non-linear positive association with CKM staging and was an independent risk factor for it (adjusted OR ≈ 3.90). Regarding mortality outcomes, MCMI was independently and positively associated with all-cause mortality risk, with a clear threshold effect (risk significantly increased when MCMI > 3.5). Its association with cardiovascular mortality was primarily mediated by diabetes mellitus (mediation proportion: 45.5%). Restricted cubic spline analysis revealed distinctly different non-linear patterns between MCMI and the two types of mortality risk. Sensitivity analyses and competing risk models further confirmed the robustness of these associations, and MCMI demonstrated significantly superior predictive ability for prognosis compared to the CMI and TyG indices. We confirmed that MCMI is a robust and independent prognostic predictor, with predictive efficacy superior to traditional metabolic indicators (CMI, TyG), providing a new quantitative tool for risk management in CKM.

First, through polynomial linear regression and ordinal logistic regression, we confirmed that MCMI has a “decelerating increase” non-linear positive association with CKM staging. This association remained an independent risk factor for stage progression after adjusting for confounders such as sex, diabetes, and hypertension (adjusted OR ≈ 3.90, p < 2e−16), suggesting that MCMI could serve as a potential metabolic marker for CKM stage progression.

The cardiometabolic index (CMI) is a composite index calculated from triglycerides (TG), high-density lipoprotein cholesterol (HDL-C), and the waist-to-height ratio (WHtR). It was initially proposed a decade ago as a direct marker for distinguishing diabetes mellitus(18). The triglyceride-glucose (TyG) index, which incorporates fasting triglyceride and fasting plasma glucose levels, is an effective surrogate biomarker for insulin resistance (IR) and is associated to varying degrees with diseases such as diabetes, cardiovascular disease, cerebrovascular disease, fatty liver, kidney disease, and reproductive system disorders(19). The modified cardiometabolic index (MCMI) is a novel indicator based on the CMI and TyG formulas, integrating multiple metabolic information including central obesity, lipids, and blood glucose. Recently, it has been found to have a non-linear positive association with non-alcoholic fatty liver disease and liver fibrosis(6).

CKM syndrome is a systemic disease arising from the pathophysiological interactions between metabolic risk factors (such as obesity and diabetes), CKD, and the cardiovascular system. It leads to multi-organ dysfunction and a high risk of adverse cardiovascular outcomes(2), with metabolic abnormalities (including obesity, insulin resistance, and hyperglycemia) being the core initiating factors. Obesity is characterized by excessive and dysfunctional adipose tissue, leading to fat accumulation. Excess lipids can be ectopically deposited in organs such as the liver, skeletal muscle, myocardium, and kidneys, disrupting insulin signaling and ultimately leading to insulin resistance(20). Insulin resistance reduces cellular glucose uptake, resulting in hyperglycemia. This hyperglycemic state can induce non-enzymatic glycation of lipids and proteins, forming advanced glycation end products (AGEs)(21). AGEs promote the accumulation and cross-linking of extracellular matrix (ECM) proteins, leading to vascular stiffness and fibrosis, which can progress to diabetic cardiomyopathy and diabetic nephropathy(22). In type 2 diabetes, insulin resistance shifts the metabolic characteristics of tissue cells from glucose metabolism towards fatty acid oxidation(4). This shift leads to excessive uptake of free fatty acids and triglycerides, generating toxic lipid metabolites and inducing mitochondrial dysfunction, thereby promoting cardiac fibrosis and diastolic dysfunction(23).

In obese states, adipocytes also secrete excessive leptin(24). Leptin not only induces glomerulosclerosis and fibrosis but also exacerbates oxidative stress in renal tubular epithelial cells—both are potential risk factors for chronic kidney disease (CKD). Furthermore, leptin contributes to hypertension by enhancing sympathetic nervous system activity and reducing nitric oxide (NO) production, thereby damaging the cardiovascular system(25). A decline in the glomerular filtration rate activates compensatory mechanisms, including the RAAS, sympathetic nervous system, and calcium-parathyroid axis, all of which adversely affect the cardiovascular system(26). Additionally, the common lipid and lipoprotein metabolism disorders in CKD patients, such as elevated total cholesterol, elevated low-density lipoprotein (LDL), and reduced high-density lipoprotein (HDL), promote inflammation and atherosclerosis, significantly increasing CVD risk(27).

MCMI integrates multiple metabolic information, including central obesity, lipids, and blood glucose, which constitute the common soil driving cardiorenal metabolic damage. Early metabolic abnormalities may have a stronger “driving effect” on organ damage. As CKM staging progresses, the influence of other factors (such as structural cardiac changes and irreversible decline in renal function) may increase. Pathological processes like the activation of the renin-angiotensin-aldosterone system (RAAS), sympathetic nervous system excitation, and irreversible organ fibrosis may dominate, thereby attenuating the marginal effect of the metabolic factors represented by MCMI. Consequently, MCMI could serve as a simple screening tool to identify populations at high risk for rapid early progression of CKM, and intervening in MCMI or the metabolic components it represents might help delay CKM stage progression.

Further Cox regression analysis confirmed an independent positive association between MCMI and all-cause mortality in the CKM population, revealing a threshold effect. Specifically, when MCMI exceeded 3.5, the risk of all-cause mortality increased significantly; the highest quartile (Q4) exhibited a 41.2% higher risk compared to the lowest quartile (Q1) (HR=1.412, P=0.024). This may represent a “tipping point” of metabolic dysregulation, beyond which systemic inflammation, oxidative stress, and endothelial dysfunction reach a critical threshold, leading to a sharp escalation in mortality risk. The identified “safety interval” of MCMI < 3.5 offers a potential reference value for establishing clinical management targets. To further characterize the dynamic nature of this risk, we employed restricted cubic spline (RCS) analysis. This revealed a monotonically increasing non-linear trend between MCMI and all-cause mortality, with a clear safety interval (HR < 1 for MCMI < 3.5). This finding complements, rather than contradicts, the threshold effect observed in the Cox model: the overall trend is upward, but the risk curve remains relatively flat at lower MCMI values, with the slope increasing sharply after the threshold is crossed. Therefore, in the clinical management of CKM syndrome, attention should be paid not only to elevated MCMI levels but also, more importantly, to values exceeding the 3.5 threshold, as this signifies the need for more intensive intervention.

However, in the Cox regression models for cardiovascular mortality in the CKM population, we found that the association was not statistically significant in the fully adjusted model but became significant after removing the diabetes covariate (HR=1.374, P=0.008). Therefore, we used mediation analysis to further confirm that diabetes mediated 45.5% of the effect of MCMI on cardiovascular mortality risk, clarifying the key mediating role of diabetes in MCMI-induced cardiovascular death. This indicates that the primary pathway through which the metabolic abnormalities represented by MCMI lead to cardiovascular death is by inducing or exacerbating diabetes, which subsequently leads to death through diabetes-related cardiovascular complications (such as atherosclerosis and cardiomyopathy). The underlying pathophysiological mechanism can be reasonably explained as follows: MCMI, as a comprehensive marker reflecting the degree of metabolic disturbance, is often accompanied by pathological changes such as insulin resistance, dysregulation of glucose and lipid metabolism, and visceral fat accumulation. These changes are precisely the core pathological basis for the development and progression of type 2 diabetes.

When the body is in a state of high MCMI for a prolonged period, insulin sensitivity decreases, and pancreatic β-cell function becomes impaired, ultimately leading to disrupted glucose homeostasis and progression to diabetes. Diabetes, as one of the most important risk factors for cardiovascular disease, can accelerate the occurrence of adverse cardiovascular outcomes through multiple pathways, including promoting the formation and progression of atherosclerosis, damaging vascular endothelial function, inducing microvascular disease, and increasing the risk of arrhythmias and heart failure, ultimately significantly elevating the risk of cardiovascular death.

This forms a clear causal chain: elevated MCMI levels → insulin resistance/impaired glucose metabolism → development of diabetes → increased cardiovascular mortality risk. The RCS curve for the association between MCMI and cardiovascular mortality risk exhibited a fluctuating “rise-fall-rise” non-linear pattern. The identification of a low-risk interval (3.0-3.5) and high-risk intervals (<2.5 or >4.0) further highlights the interval-specific nature of the MCMI risk effect. The lowest risk interval (3.0-3.5) may represent a state of “metabolic adaptation,” while the increased risk associated with extremely high (>4.0) and extremely low (<2.5) MCMI may reflect severe metabolic syndrome and the risk of malnutrition, sarcopenia, or underlying chronic wasting diseases, respectively.

Subgroup analyses showed that the association between MCMI and mortality risk was more pronounced in middle-aged and older adults, males, and individuals not using antihypertensive medications or statins. However, interaction analyses did not reveal significant heterogeneity, supporting the robustness of MCMI as a prognostic risk factor. The stronger association between MCMI and mortality risk in the untreated population suggests that medications (antihypertensives, statins) may partially attenuate the metabolic risk represented by MCMI, further demonstrating the effectiveness of these drugs and indicating a higher predictive value of MCMI in untreated populations. To further exclude potential bias and verify the reliability of the results, we performed sensitivity analyses by excluding specific populations, altering the racial composition, and adjusting for different covariate models; the results remained robust, greatly enhancing the credibility of the conclusions. More importantly, E-value analysis indicated that 推翻 this significant independent association would require hypothesizing the existence of an unmeasured confounder of moderate to strong strength. Based on current biological knowledge and clinical practice, the existence of such strong confounders not included in the model is unlikely. Therefore, although observational studies cannot completely exclude residual confounding, the comprehensive analysis suggests that MCMI is likely an independent risk marker for all-cause mortality.

Notably, the results from the competing risk model corrected the limitations of the traditional cause-specific Cox model. After accounting for competing events of non-CVD death, the strength of the association between MCMI and CVD death increased substantially, demonstrating a clear dose-response relationship (Q4 vs. Q1 sHR=3.25, trend P<0.001). This indicates that ignoring competing risks severely underestimates the true effect of MCMI on CVD death. Neither traditional all-cause mortality analysis (HR=1.43) nor the cause-specific Cox model ignoring competing risks fully revealed the strong association between MCMI and CVD death (with sHR reaching as high as 3.25). This discrepancy clearly suggests that non-CVD death, as a competing event, ‘masks’ the true risk of MCMI for CVD death. Therefore, MCMI likely influences prognosis by specifically exacerbating cardiovascular pathophysiological processes (such as atherosclerosis, myocardial fibrosis, etc.), rather than non-specifically increasing the risk of various causes of death. Future research should focus on exploring the specific biological mechanisms through which MCMI affects cardiovascular health. Finally, the comparison of predictive values showed that MCMI’s independent predictive ability for prognosis in the CKM population was significantly superior to that of CMI and TyG, providing a more precise metabolic indicator for clinical risk stratification.

The strengths of our study include: (1) We utilized the latest PREVENT equation recommended by the American Heart Association (AHA) to predict subclinical cardiovascular disease (CVD) and employed the new race-free estimated glomerular filtration rate (eGFR) equation to calculate eGFR, making the CKM staging more precise. (2) Based on a large-scale, prospective community-based cohort, we systematically evaluated the predictive role of MCMI across the entire spectrum of CKM. (3) We employed advanced analytical strategies, such as RCS curves to reveal non-linear associations, competing risk models to correct for estimation bias, and mediation analysis to preliminarily elucidate biological pathways. (4) We also used a series of rigorous sensitivity analyses and E-value assessments to jointly confirm the robustness of the study conclusions. (5) We not only discovered the independent predictive value of MCMI and its clinical cut-off value (>3.5) but also confirmed its superior predictive efficacy over traditional metabolic indicators through head-to-head comparisons; these findings have clear potential for clinical translation.

Simultaneously, our study has limitations: (1) The observational study design precludes the determination of causality between MCMI and outcomes, only allowing for the suggestion of associations. (2) Despite comprehensive adjustment for covariates, there may be unmeasured confounding factors (e.g., dietary details, genetic factors). (3) Our study is based on a specific community-based cohort (NHANES), and caution is needed when extrapolating the conclusions to other racial or regional populations. (4) This study primarily focused on epidemiological associations, with limited exploration of the underlying biological mechanisms (e.g., inflammation, hormone levels).

In conclusion, our study demonstrates that MCMI is an independent and robust predictor of CKM stage progression and all-cause mortality risk. Its association with cardiovascular mortality is primarily mediated by diabetes, and it exhibits complex non-linear threshold effects. Future prospective interventional studies are needed to verify whether reducing MCMI can improve CKM prognosis and to validate the predictive efficacy of MCMI in more diverse populations.

## 5. Conclusion

In summary, this study, based on a large-scale community-based cohort of patients with Cardiovascular-Kidney-Metabolic (CKM) syndrome, is the first to systematically reveal the independent predictive value of the modified cardiometabolic index (MCMI) across the entire spectrum of CKM syndrome. The study confirms that MCMI is not only an independent risk factor for CKM stage progression (exhibiting a “decelerating increase” non-linear association) but also a robust predictor of all-cause mortality risk, establishing MCMI > 3.5 as a potential clinical cut-off value for risk management. Importantly, we found that diabetes mediated 45.5% of the MCMI-associated cardiovascular mortality risk, elucidating the core pathway of “MCMI → diabetes → cardiovascular death.” Furthermore, competing risk models revealed that failing to account for non-cardiovascular death events would substantially underestimate the true effect of MCMI (sHR reaching 3.25). Additionally, MCMI demonstrated significantly superior predictive performance for prognosis in the CKM population compared to traditional indicators (CMI, TyG), providing a superior quantitative tool for precise clinical risk stratification. This study transcends the limitations of applying MCMI to single-organ diseases, offering new evidence-based support for the early identification, risk stratification, and individualized intervention of CKM syndrome. Future prospective interventional studies and cross-ethnic cohort validations are needed to facilitate the clinical translation and application of MCMI.

## Abbreviations

All-cause mortality Antihypertensive use: All-cause mortality Antihypertensive medication use
BMI: Body Mass Index
CKM: Cardiovascular-Kidney-Metabolic syndrome
CVD death: Cardiovascular disease death
HbA1c: Glycated Hemoglobin
HDL-C: High-Density Lipoprotein Cholesterol
LDL-C: Low-Density Lipoprotein Cholesterol
MCMI: Modified Cardiometabolic index
mDBP: Mean Diastolic Blood Pressure
Mets: Metabolic syndrome
mSBP: Mean Systolic Blood Pressure
SUA: Serum Uric Acid
TC: Total Cholesterol
TG: Triglycerides
UACR: Urine Albumin-to-Creatinine Ratio
WC: Waist Circumference

## Acknowledgements

We extend our sincere appreciation to every participant involved in the NHANES study and the entire project team.

## Author contributions

Qin Yuqi was responsible for study design, conceptualization, data collection, statistical analysis, result interpretation, as well as drafting and revising the manuscript. Yan Yibo was responsible for study design, reviewing and revising the manuscript. All authors read and approved the manuscript.

## Data availability

The datasets generated and analyzed during this research can be retrieved from the NHANES database (https://wwwn.cdc.gov/nchs/nhanes/default.aspx).

## Declarations

### Approval from the ethics committee and consent for participation

The research protocol of the NHANES study obtained approval from the Ethics Review Board of the National Center for Health Statistics. Prior to their participation, all subjects signed a written informed consent form.

